# LungAI: An AI Biomarker that Refines Lung-RADS v2022 Triage of Screen-Detected Pulmonary Nodules

**DOI:** 10.64898/2026.11.09.26367662

**Authors:** Huzaifa Suri, Yakov Keselman, Lance Legel, Jonathan Govette, Bradley Mullen, Nirmal Choradia, Thamer Sartawi, Fereidoun Abtin, Drew Moghanaki, Carey Thomson, David Yankelevitz, Yuri Ahuja

## Abstract

**Background:** The Lung-RADS algorithm standardizes the management of screen-detected pulmonary nodules, but its risk assessment is driven chiefly by nodule size and attenuation. This misallocates a meaningful fraction of patients: a nontrivial share of small malignancies receive 12-month surveillance while large benign nodules proceed to invasive workup. Artificial intelligence (AI) may supply risk information independent of the imaging features Lung-RADS already uses.

**Methods:** We validated LungAI, an investigational deep learning nodule malignancy classifier not yet cleared by the US Food and Drug Administration (FDA), on the held-out NLST test partition of Ardila et al. (2,747 low-dose CT examinations from 1,244 patients with ≥1 solid or part-solid 4-30 mm nodule; 89 pathologically confirmed cancers, prevalence 3.2%). We compared discrimination against Sybil, mean nodule diameter, and estimated nodule volume. A board-certified radiologist assigned Lung-RADS v2022 categories to a positives-enriched random subset (226 eligible examinations; 78 cancers), projected to the full cohort by contingency weighting. A per-category reclassification algorithm shifted patients along the existing Lung-RADS management ladder using a-priori score cutpoints. We evaluated a separately locked follow-up configuration on examinations with a prior scan.

**Results:** LungAI achieved AUROC 0.92 (95% CI 0.88-0.95) and AUPRC 0.47 (0.35-0.61), outperforming Sybil (ΔAUROC 0.07; ΔAUPRC 0.18), diameter (ΔAUROC 0.14; ΔAUPRC 0.34), and volume (ΔAUROC 0.14; ΔAUPRC 0.38; all P<0.001). Against Lung-RADS v2022 scored ordinally on the weighted annotated subset, LungAI improved AUPRC (ΔAUPRC 0.31, P=0.003) but not AUROC (ΔAUROC 0.03, P=0.20). LungAI added discriminative information independent of diameter (likelihood-ratio χ^2^=161, P<10^−36^; ΔAUROC 0.10) and, on follow-up examinations, independent of delta diameter, volume doubling time, and new-nodule status (χ^2^=79, P<10^−18^). Integrated with Lung-RADS, LungAI reduced cancers on the slowest (12-month rescan) pathway by 46% (3.1→1.6 per 1,000 examinations) and benign biopsy referrals by 20% (32.7→26.2 per 1,000 examinations) at approximately unchanged total biopsy volume (51.0→50.1 per 1,000). Assuming full adherence to recommended follow-up, the combined approach yielded 25 months of net diagnostic lead time, an estimated net cost saving of ∼$2,200, and 6.5 fewer benign biopsies per 1,000 examinations; accounting for published adherence rates, it yielded 1.6 additional cancer diagnoses within 1 year per 1,000 examinations.

**Conclusions:** LungAI contributes malignancy-risk information orthogonal to the size- and growth-based features underlying Lung-RADS and improves the allocation of screening patients across the existing management spectrum while potentially decreasing invasive procedures for benign lesions. Multi-site, multi-geography validation is required to demonstrate practical generalizability.

## Introduction

Lung cancer remains the leading cause of cancer mortality worldwide, responsible for an estimated 1.8 million deaths in 2022 [1]. Prognosis is strongly stage-dependent: five-year survival is 68-92% for stage I disease but falls below 10% at stage IV [2]. Because a substantial fraction of tumors are still diagnosed after regional or distant spread, strategies that shift detection to earlier stages have considerable potential to reduce population mortality.

Randomized trials have established that annual low-dose computed tomography (LDCT) meaningfully reduces lung cancer mortality, by 20% in the National Lung Screening Trial (NLST) and by 24-33% in the NELSON trial [3,4]. In 2021, the US Preventive Services Task Force broadened its recommendation to adults aged 50-80 years with a ≥20 pack-year history, expanding eligibility by approximately 6.4 million people [5]. As the screened population grows, the discriminative accuracy of nodule risk assessment becomes increasingly important to contain false positives without sacrificing sensitivity.

The American College of Radiology’s Lung-RADS system, introduced in 2014 and refined in version 2022 [6], standardized nodule reporting and management. However, its risk assessment relies mainly on nodule diameter and attenuation; secondary features such as margins and spiculation are assessed subjectively, with inter-reader disagreement in 29% of reading pairs [7]. The consequences run in both directions. The positive predictive value of Lung-RADS 4B is only 22–37% [8], and transthoracic needle biopsy carries meaningful complication risk, including pneumothorax in approximately 15% of procedures (6.6% requiring chest-tube placement) and hemorrhage in 1% [9]. Conversely, malignant nodules assigned to low categories wait up to a year for their next evaluation, risking meaningful disease progression and prognostic consequence. Clinical risk calculators, including the Mayo [10] and Brock [11] models, and NELSON’s volumetric approach, add structured probability estimation but rely largely on the same morphological inputs.

AI-based nodule classification has emerged along two lines. Per-nodule characterization tools, including Optellum’s LCP-CNN [12] and RevealDx’s RevealAI-Lung [13], hold clearance for incidentally detected nodules. Scan-level forecasting models such as Sybil [14] predict future cancer risk but are not FDA cleared. To our knowledge, Median Technologies’ eyonis LCS holds the only FDA clearance with a screening-specific indication [15]. We propose that the value of any such tool is best understood through the concept of an AI biomarker: an algorithmic measurement whose worth lies precisely in the information it contributes *orthogonal* to the variables already driving existing estimators.

Here we validate LungAI, a deep learning malignancy classifier producing per-nodule and per-examination risk scores, on a held-out NLST partition identical to the published test split of Sybil and Ardila et al. [14,16]. We deliberately position LungAI as an adjunct to, not a replacement for, Lung-RADS v2022. The reclassification algorithm we propose preserves the familiar management ladder (12-month, 6-month, 3-month, biopsy) and shifts patients within it, requiring no change to radiology workflow or reporting infrastructure. We test four propositions: (i) LungAI outperforms Sybil and size-based measures on discrimination; (ii) it retains discriminative value after adjustment for diameter – and, on follow-up examinations, after adjustment for interval growth – enabling stratification within Lung-RADS categories; (iii) its score strata map monotonically onto clinically meaningful risk tiers; and (iv) performance is stable across patient and technical subgroups.

## Methods

### Study design and oversight

This retrospective diagnostic-accuracy study used de-identified NLST data and is reported per STARD 2015 [17] (flow in Supplementary Figure S1). We locked the LungAI model weights, follow-up risk adjustment configuration, and all reclassification rules before evaluating the held-out test partition. This secondary analysis used de-identified NLST data obtained from the NCI Cancer Data Access System under an approved data-use agreement; no investigator had access to identifiable information, and institutional review board review was therefore not required.

### Cohort

The evaluation cohort was the NLST held-out test partition defined by Ardila et al. [16] and used as the published test split of Sybil [14]; it was never used in either LungAI or Sybil development. NLST enrolled high-risk current and former smokers aged 55-74 between 2002 and 2004 with annual LDCT screening and pathologically confirmed outcomes [3].

Within this partition, all LDCT examinations with ≥1 nodule detected by Coreline Soft’s FDA-cleared aView LCS nodule-detection software [18] were considered (4,872 examinations, 1,817 patients, 128 cancers). Examinations were excluded if the cancer-associated nodule was not visible on the index LDCT (164 examinations), slice thickness exceeded 2.5 mm (257 examinations, 7 cancers), or no solid or part-solid nodule measured 4-30 mm (1,704 examinations, 32 cancers). We excluded ground-glass nodules a priori because ground-glass malignancies are often indolent and managed with surveillance rather than resection, making a malignancy score less decision-relevant. The analysis set comprised 2,747 examinations from 1,244 patients (89 cancers, 3.2%). Notably, only 7 examinations (0.3%) included part-solid nodules and none were malignant; the analyzed cohort is therefore de facto solid-nodule-only.

### Index test

LungAI is a locked, commercial-grade, deep learning computer-aided diagnosis (CADx) model that analyzes LDCT volumes and returns a malignancy score (0.0 – 10.0, in increments of 0.1) for each detected nodule and an aggregate examination-level score defined as the maximum across nodule scores. The examination-level maximum reflects the clinical decision unit: triage is determined by the most suspicious nodule while per-nodule scores preserve interpretability and localization. LungAI scores are not probabilities; they are unitless scores with absolute-risk interpretation provided empirically through observed malignant fractions per score stratum (Table S1). This non-probability score convention is common among cleared CADx devices [12]. The displayed scale is anchored to fixed operating points estimated on the NLST validation partition: integer scores 1.0 through 9.0 correspond, respectively, to thresholds operating at 95%, 90%, and 85% sensitivity and at 90%, 95%, 97%, 98%, 99%, and 99.5% specificity. A threshold at a given score therefore carries a locked operating interpretation; for example, a rule flagging examinations with scores at or above 2.0 operates at approximately 90% sensitivity in the validation population. This anchor mapping was frozen with the model before any evaluation of the held-out test partition. LungAI is investigational: a 510(k) premarket notification is pending with the US Food and Drug Administration, and the device is not cleared or available for sale in the United States.

Two locked configurations were evaluated. The standalone configuration, hereafter referenced as LungAI, operates on a single examination. The follow-up configuration, hereafter referenced as LungAI-FU, is a logistic regression on three inputs per nodule: the standalone-model raw output on the current examination, the nodule’s inverse volume doubling time relative to the matched prior nodule, and an indicator for tracked versus new nodules. Inverse volume doubling time was expressed as log_2_(Vcurrent/Vprior) / Δt (doublings per year). Untracked nodules carry no growth term, and tracked nodules whose prior detection has no measurement receive the tracking indicator with growth term set to zero (458 nodules, all benign). LungAI-FU is applicable when a prior examination exists, mirroring standard clinical practice in which screening studies are interpreted against priors. Nodule measurement on both examinations and inter-examination nodule matching were performed using Coreline’s aView LCS. We fit and cross-validated fusion coefficients on the NLST validation partition, leaving the test partition untouched before evaluation; we report locked coefficients as odds ratios in Table S2. The follow-up analysis set comprised all post-baseline examinations (n=1,899; 45 cancers), coinciding with the screening-round stratification of Supplementary Figure S2.

### Comparators

#### Lung-RADS v2022

NLST pre-dates Lung-RADS. Lung-RADS categories were therefore assigned retrospectively by a board-certified radiologist according to ACR v2022 rules, blinded to LungAI outputs and to outcomes, and without prior-scan information.

Annotation covered a positive-enriched random subset of 256 examinations, of which 226 met analysis-set eligibility (78 cancers, 148 benign; sampling fractions 87.6% of cancers, 5.6% of benign examinations). All Lung-RADS-dependent quantities were projected to the full analysis set by contingency weighting (w+ = 89/78 ≈ 1.14; 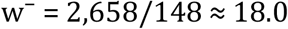 Table S3). We evaluated Lung-RADS’s discriminative performance on the annotated subset with contingency weighting and performed inference on ΔAUROC and ΔAUPRC by patient-clustered bootstrap. At the 3/4A decision boundary, we measured weighted sensitivity at 81.2% and specificity at 85.1%.

#### Sybil

The publicly released Sybil model [14] was applied to identical examinations using its one-year risk score, with no nodule-coordinate input as designed. Comparison between LungAI and Sybil was only performed at the examination level, where both produce a score. We selected the one-year horizon as the most favorable to Sybil.

#### Size measures

We studied mean nodule diameter and nodule volume as standalone discriminators. On follow-up examinations, interval change in diameter (Δdiameter) and volume (Δvolume) were used as comparators. These measures are defined only for tracked nodules with a measured prior and were evaluated on the 963 follow-up examinations with at least one such nodule (27 cancers).

### Reference standard

An examination was labeled as malignant if a pathologically confirmed primary lung cancer was recorded for the corresponding nodule at any time within the NLST follow-up window. An examination was labeled as benign if the trial read was negative with no cancer diagnosis during follow-up.

### Reclassification algorithm

The combined algorithm preserves the Lung-RADS management ladder and shifts patients within it by thresholding the displayed LungAI score: up-triage advances management by one rung (12-month LDCT → 6-month LDCT → 3-month LDCT/PET-CT → biopsy referral) and down-triage retreats by one. We evaluated two reclassification schemata, both were specified before evaluation, and we tested no other cutpoint configurations.

#### Schema A (primary): per-category cutpoints matched to Lung-RADS’s own sensitivity profile

Round cutpoints were fixed a priori within each Lung-RADS category. Up-triage cutpoints were fixed at 2.0 (empiric sensitivity ∼90%) within Lung-RADS 2, 4.0 (empiric specificity ∼90%) within Lung-RADS 3, and 6.0 (empiric specificity ∼97%) within Lung-RADS 4A, roughly corresponding to published metrics for Lung-RADS 3, 4A, and 4B respectively. We chose these cutpoints to match our empiric estimate of Lung-RADS v2022’s own performance at the 2/3, 3/4A, and 4A/4B decision boundaries on the annotated NLST subset, and consistent with published estimates for Lung-RADS version 1.0 (84.9% baseline sensitivity at the category-≥3 positivity threshold) [19]. Down-triage cutpoints were fixed at 1.0 in Lung-RADS 3 (empiric sensitivity ∼95%), 1.5 in Lung-RADS 4A (arithmetic midpoint of 1.0 and 2.0), and 2.0 in Lung-RADS 4B/4X (empiric sensitivity ∼90%). We chose these cutpoints to be highly conservative in down-triaging, reflecting the logic that an erroneous down-triage decision is significantly more costly than an erroneous up-triage one. The cutpoints were pre-selected and applied once without modification. Achieved within-category sensitivities and specificities are reported in Results and were not used to revise the cutpoints post-hoc.

#### Schema B (sensitivity analysis): a single operating point

Up-triage at a single displayed score of 4.0 across all categories, with down-triage cutpoints unchanged. The cutpoint 4.0 (empiric specificity ∼90%) was chosen as a round clinical operating point approximating the maximum-Youden threshold derived on the NLST validation partition. Results are in Supplementary Figure S3.

#### Robustness

A threshold-perturbation analysis jointly scaled the operating thresholds of all six Schema A cutpoints across a four-fold range (×0.50 to ×2.00) and recomputed the principal reclassification metrics (Table S4).

### Statistical analysis

Our primary endpoint was examination-level AUROC, which we compared against each comparator risk score with the paired DeLong test [20]. AUPRC differences were tested by paired patient-clustered bootstrap (2,000 replicates). All confidence intervals were obtained by the same patient-clustered bootstrap. We additionally assessed model discrimination at the nodule level, wherein each cancer examination contributed one nodule (the diagnosed target nodule) and each benign examination contributed all nodules meeting inclusion criteria. Since including multiple nodules per examination violates the independence assumption of the DeLong test, we used patient-clustered bootstrap for nodule-level inference. Sybil, which produces no nodule-level output, was omitted from nodule-level analysis (Table S5; Supplementary Figure S4).

Orthogonality was tested with nested logistic models and likelihood-ratio tests: at baseline, malignancy ∼ restricted cubic spline of diameter (3 df) ± LungAI; on follow-up examinations, M0 = spline(diameter) + volume doubling time + new-nodule indicator versus M1 = M0 + LungAI standalone (6.4 events per parameter). Reclassification was tested by McNemar’s exact test separately among cancers and benign examinations on the annotated subset, with weighted projection for cohort-level counts, and summarized by weighted categorical NRI [21]. Stage at diagnosis was taken from the NLST cancer data and assigned to every examination of the patient. Among annotated cancers we tabulated stage by direction of the LungAI-based management change (Table S6). Subgroup heterogeneity used Cochran’s Q with I^2^. Decision curve analysis [22] compared net benefit of Lung-RADS alone versus the combined algorithm over threshold probabilities of 1-20%, with stratum-level observed malignant fractions from the weighted cross-tabulation as probability inputs. Since Lung-RADS categories exist only on the annotated evaluation subset, these risk estimates are necessarily in-sample, and the decision-curve comparison should be read as descriptive rather than out-of-sample (Supplementary Figure S5).

The clinical-economic analysis is described in Figure 5. We use Medicare-anchored unit costs (LDCT$300; CT-guided biopsy$4,000; Table S7) [23], lead-time accounting netting delayed down-triaged cancers, and a real-world scenario applying published “defined adherence” rates per Lung-RADS v2022 category (Table S8). Adherence rates were taken as the unweighted mean of four studies [24–27] identified in the meta-analysis of Lin et al. [28], under the assumption that patients on a 12-month pathway have, by definition, no opportunity for diagnosis within 1 year. All hypothesis tests were two-sided with alpha set to 0.05. Analyses were performed in Python 3.9.2 (numpy 1.26.4, pandas 2.3.2, scipy 1.13.1, scikit-learn 1.5.2, statsmodels 0.14.6).

## Results

### Cohort

The analysis set comprised 2,747 examinations (1,244 patients; 89 cancers, 3.2%); characteristics are in Table S9. Median age was 61 years (IQR 58-66), 62% were male, and median smoking exposure was 49.5 pack-years. Median maximal nodule diameter was 6.3 mm overall and 12.8 mm among cancers. The cohort was 93.2% White, reflecting NLST enrollment.

### Discrimination (standalone configuration)

LungAI achieved an AUROC of 0.92 (95% CI 0.88-0.95) and an AUPRC of 0.47 (0.35-0.61) (Figure 1; Table S10). It outperformed Sybil (AUROC 0.85, ΔAUROC 0.07, 95% CI 0.04-0.09, AUPRC 0.25, ΔAUPRC 0.18, 95% CI 0.11-0.25), mean diameter (AUROC 0.78, ΔAUROC 0.14, 95% CI 0.08-0.19, AUPRC 0.13, ΔAUPRC 0.34, 95% CI 0.22-0.47), and volume (AUROC 0.79, ΔAUROC 0.14, 95% CI 0.09-0.20, AUPRC 0.12, ΔAUPRC 0.38, 95% CI 0.25-0.50) on this cohort. The AUPRC separation – nearly doubling precision-recall performance relative to the best comparator at 3.2% prevalence – is the most clinically salient difference. Lung-RADS v2022 itself, scored ordinally on the weighted annotated subset, achieved AUROC 0.89 (95% CI 0.82-0.94) and AUPRC 0.25 (95% CI 0.15-0.47). LungAI’s AUROC advantage over Lung-RADS was not significant (ΔAUROC 0.03, 95% CI -0.04-0.10, P=0.20), while its AUPRC advantage was significant (ΔAUPRC 0.31, 95% CI 0.10-0.50, P=0.003). Nodule-level discrimination was concordant: the per-nodule score achieved AUROC 0.93 (0.89-0.96) and AUPRC 0.37 (0.23-0.57) at 1.5% nodule-level prevalence, exceeding diameter and volume on both metrics (all P≤0.001; Table S5; Supplementary Figure S4).

**Figure 1.**
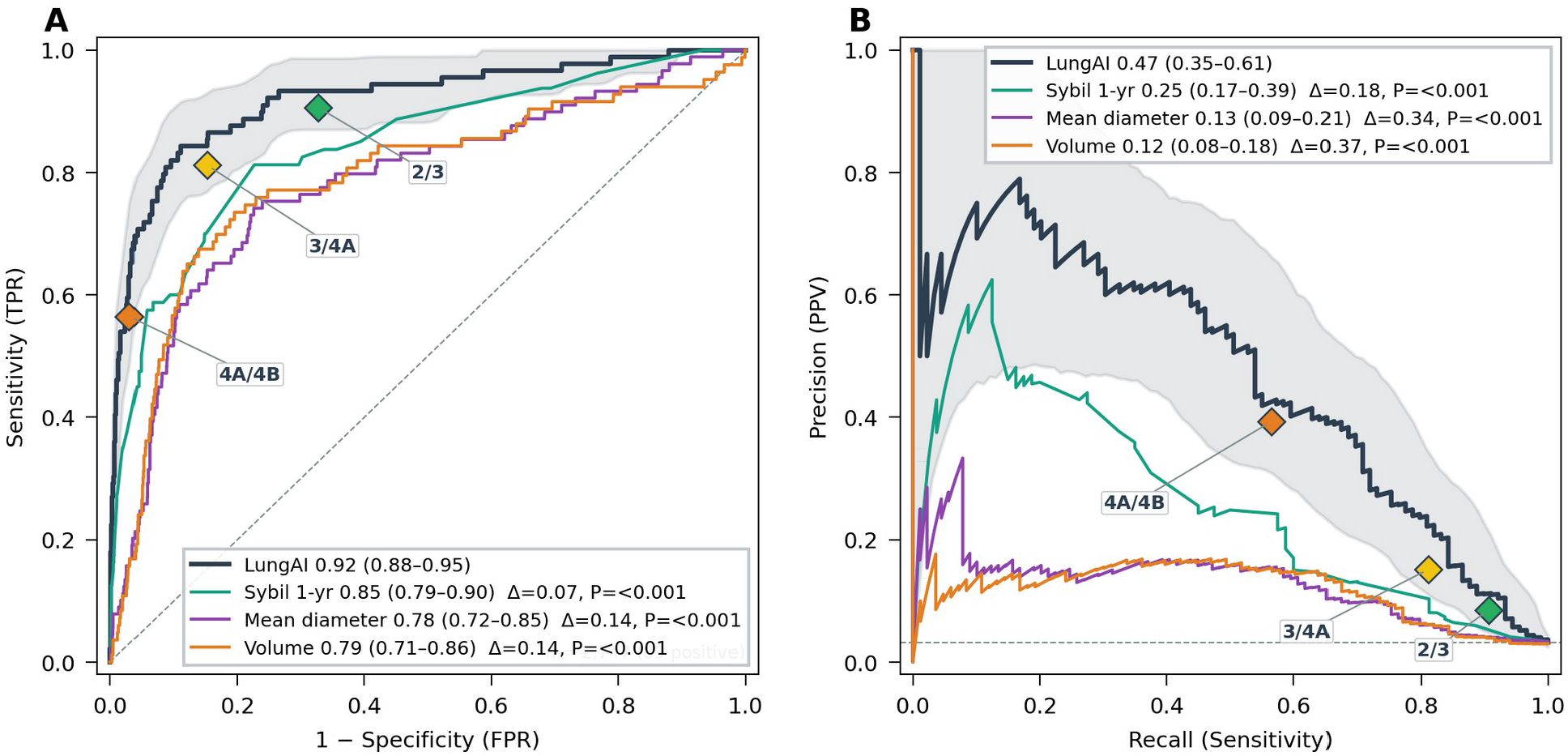
Standalone model discrimination without prior scan information. Examination-level (A) receiver operating curves and (B) precision-recall curves for LungAI, Sybil (1-year), mean diameter, and estimated volume, with weighted Lung-RADS category operating points overlaid; ΔAUROC and DeLong P in legend.

### Orthogonality to nodule size

Adding LungAI to a flexible diameter model increased AUROC by 0.10 (0.06-0.18; likelihood-ratio χ^2^=161, P<10^−36^). This suggests that LungAI is not simply recapitulating nodule size.

### Follow-up configuration

Among 1,899 follow-up examinations (45 cancers), the follow-up configuration achieved AUROC 0.91 (0.83-0.97) and AUPRC 0.60 (0.45-0.74) (Figure 2), exceeding Sybil (ΔAUROC 0.06, P<0.001), Δdiameter (ΔAUROC 0.11, P=0.009), and Δvolume (ΔAUROC 0.09, P=0.021). Its advantage over the baseline configuration on the same examinations was confined to the precision-recall tradeoff (AUPRC 0.60 vs 0.41, ΔAUPRC 0.18, P<0.001); the AUROC gain was not significant (ΔAUROC 0.01, DeLong P=0.13, bootstrap P=0.078). This suggests that incorporating interval growth primarily sharpens the ranking of true cancers among high-scoring nodules. Notably, against a model of size, interval growth, and nodule novelty, LungAI remained decisively informative (χ^2^=79.4, P<10^−18^, model AUROC 0.81→0.87). This suggests that the morphological signal it captures is independently predictive even where growth information is available.

**Figure 2.**
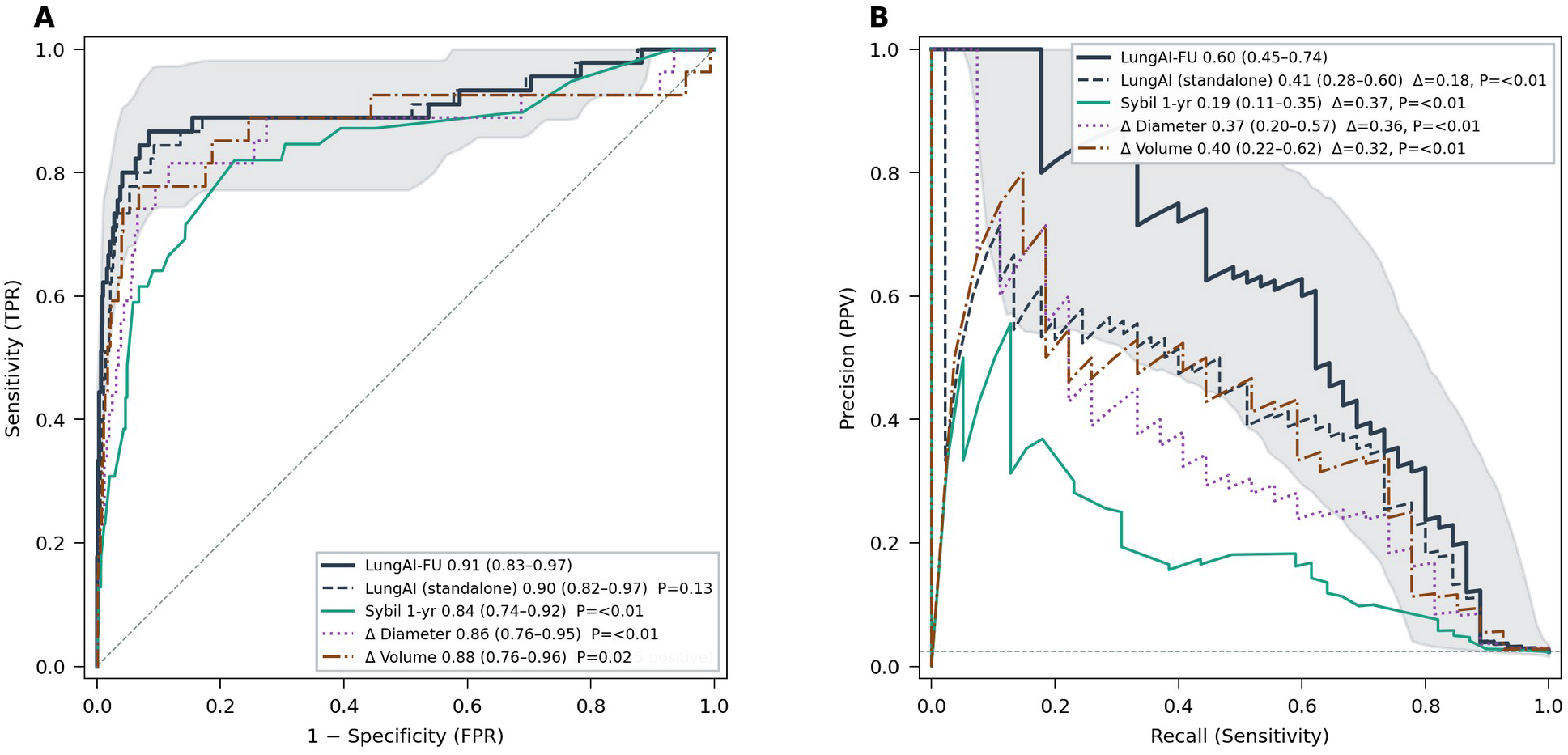
Follow-up scan discrimination (n=1,899; 45 cancers). (A) ROC and (B) PRC curves for the follow-up versus standalone configuration, Sybil (1-yr), Δdiameter, Δvolume; DeLong P in legend. Δdiameter and Δvolume are evaluated on the 963 examinations with a tracked prior nodule (27 cancers).

### Within-category stratification and reclassification

Across every Lung-RADS category, LungAI strata separated observed cancer cases substantially (Table S1; Figure 3). Within Lung-RADS 2, 0.2% of cases below a LungAI score of 2.0 were malignant versus 1.3% at or above; within Lung-RADS 3, 0.9% below a score of were malignant versus 16.7% at or above 4.0; within 4A, 1.0% below 1.5 were malignant versus 46.2% at or above 6.0; within 4B/4X, 0% below 2.0 were malignant versus 48.3% at or above it. Achieved within-category up-triage sensitivities were 78%, 50%, and 68%, capturing a meaningful proportion of cases under-triaged by Lung-RADS v2022.

**Figure 3.**
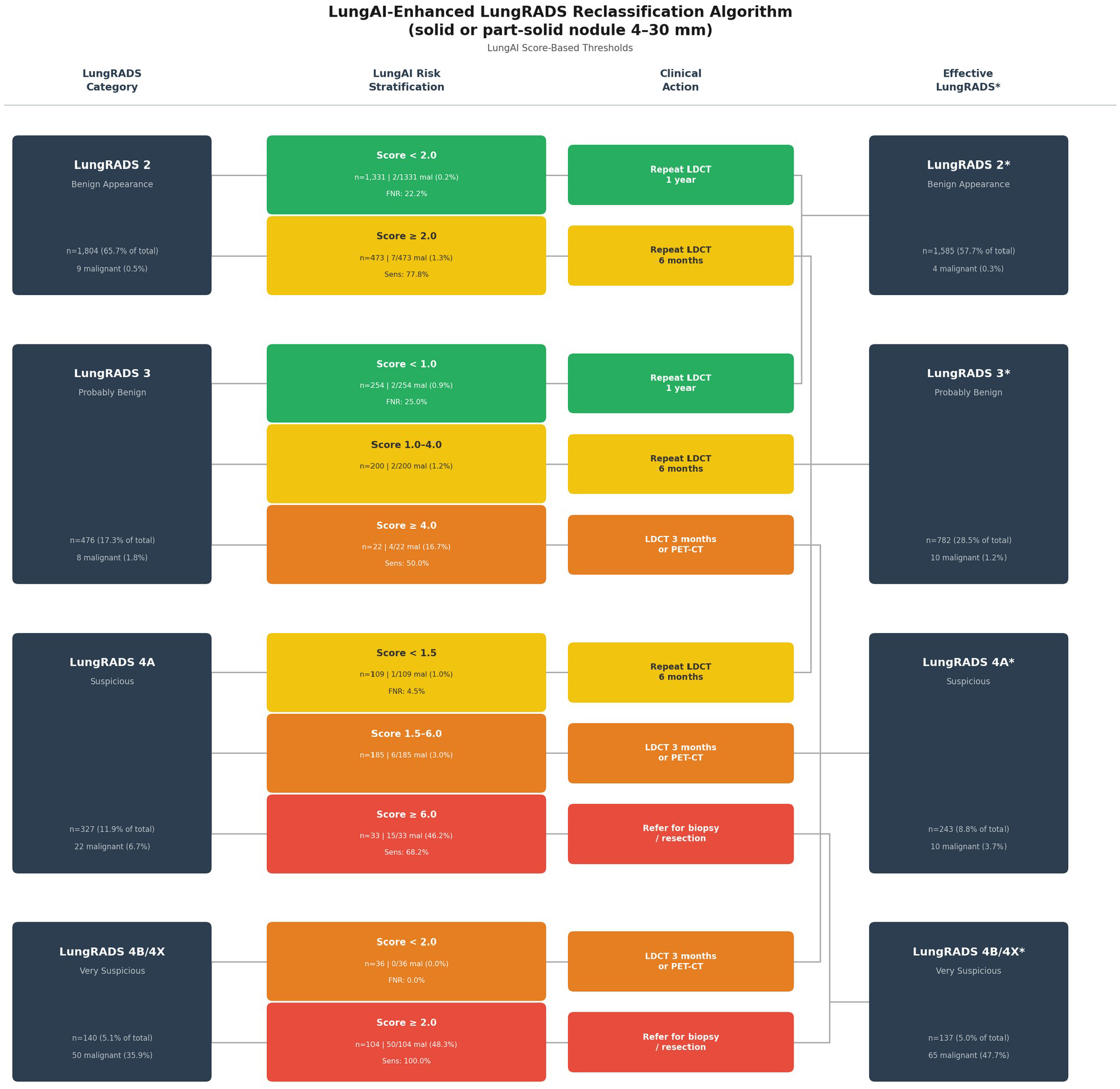
Schema A reclassification threshold tree using pre-defined per-category LungAI score strata. Includes contingency-weighted counts, observed malignant fractions, arm-level weighted sensitivity and false-negative rates, assigned management, and resulting effective Lung-RADS categories. The Schema B tree – a single up-triage operating point at a displayed score of 4.0, approximating the validation-partition maximum-Youden threshold – is shown in Supplementary Figure S3.

Applying the combined algorithm (Figure 4), cancers on the slowest, 12-month pathway fell by 46% (3.1→1.6 per 1,000 examinations) and cancers referred directly for tissue diagnosis rose 31% (18.3→23.9 per 1,000). On the other hand, benign biopsy referrals fell 20% (32.7→26.2 per 1,000) without meaningfully affecting total biopsy volume (51.0→50.1 per 1,000). Among cancers, up-triage dominated down-triage 20 to 3 (McNemar P<0.001). Among benign examinations, reclassification was approximately balanced (28 up-triaged versus 22 down-triaged, P=0.480), while invasive workup of benign patients nonetheless declined. The event NRI was +0.25 (95% CI 0.13-0.37) and the non-event NRI was -0.04 (95% CI -0.13-0.05), yielding an overall NRI of +0.20 (95% CI 0.04-0.36) (Table S11). The net reclassification benefit is positive even on the blunt overall metric, which weights a shift from 12-to 6-month imaging the same as a biopsy referral.

**Figure 4.**
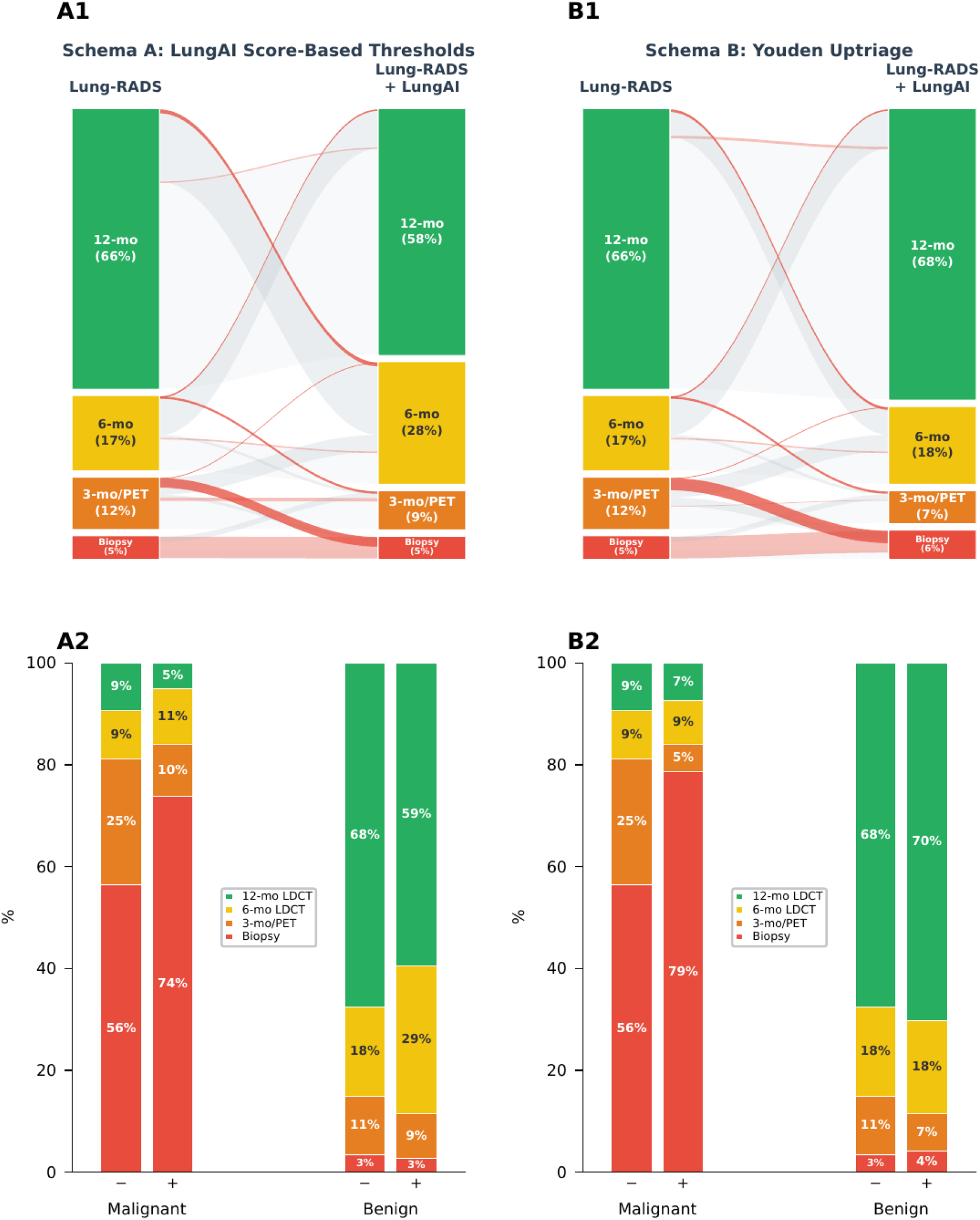
Management redistribution under per-category thresholds (Schema A) and a single max Youden operating point (Schema B). (A1, B1) Flow from Lung-RADS–assigned to combined-algorithm management under Schema A and Schema B, respectively. (A2, B2) Management distribution before versus after, separately for malignant and benign examinations, under each schema.

These findings were robust to choice of clinical operating point. Across joint scaling of all six of the above decision thresholds over a four-fold range, the event NRI remained positive throughout (+0.16 to +0.27), cancers remaining on the 12-month pathway stayed at 1.6 per 1,000 examinations (versus 3.1 under Lung-RADS alone), and net lead time remained strongly positive (18-28 months). The direction of effect is stable across the threshold range, with magnitude varying as expected (Table S4).

Stage at diagnosis, tabulated by direction of reclassification among annotated cancers (Table S6), was distributed similarly across groups: 80.0% of up-triaged cancers with known stage were stage I, versus 75.9% of unchanged cancers (exploratory Fisher exact for stage II+ versus stage I, up-triaged versus all others: OR 0.70, P=0.77). Two of the three down-triaged cancers were stage II+. Because stage reflects the care delivered in NLST, which predates modern nodule management, these analyses are purely descriptive.

### Clinical and economic trade-off

Per 1,000 nodule-positive examinations, assuming 100% adherence to LDCT rescan recommendations, the algorithm resulted in 31.7 months of gross diagnostic lead time among accelerated cancer diagnoses, less 6.4 months of delayed diagnosis, for a net lead time of 25.2 months. This comes at a net saving of approximately$2,200, with 6.5 fewer net benign biopsies (Figure 5). Applying published transthoracic-biopsy complication rates [9] to the avoided benign biopsies, this corresponds to approximately 1.0 fewer pneumothoraces, 0.4 fewer chest-tube placements, and 0.07 fewer biopsy-related hemorrhages per 1,000 nodule-positive examinations. Assuming published follow-up adherence rates (Figure 5B) [24–28], 24.9 of 32.4 cancers per 1,000 nodule-positive examinations would be diagnosed within 1 year using Lung-RADS alone versus 26.5 of 32.4 using LungAI, netting 1.6 additional timely diagnoses while at a net cost saving. Against the stage gradient of lung cancer survival [2], months of lead time at the stage I/II boundary are a plausible mechanism by which triage reallocation could translate into mortality benefit.

**Figure 5.**
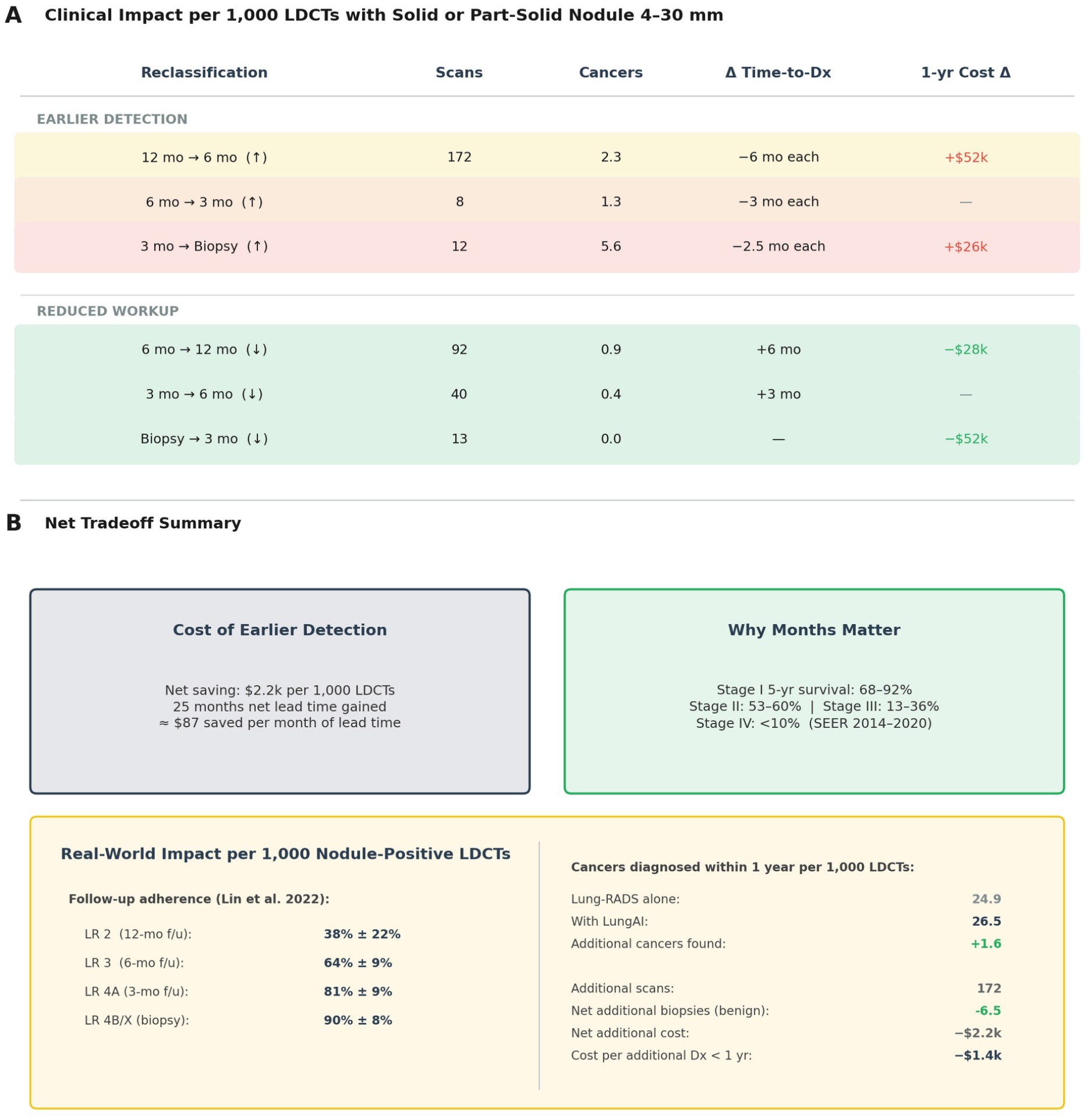
Clinical-economic trade-off per 1,000 nodule-positive examinations. (A) Reclassification transitions with lead-time and cost deltas. Net lead time was 25 months at a net saving of ∼$2,200 (approximately$87 saved per month of net lead time). (B) Real-world scenario under published adherence rates; 1.6 additional diagnoses were made within 1 year, achieved at a net cost saving. Patients on a 12-month pathway are assumed to have no opportunity for diagnosis within 1 year.

### Subgroup stability

AUROC was consistent across sex, age, scanner manufacturer, screening round, slice thickness, pack-year smoking history, and diameter bands (all Cochran’s Q P≥0.058; Supplementary Figure S2; Table S12). Cancer stage showed nominal heterogeneity (stage I versus benign AUROC 0.95, 95% CI 0.92-0.98; stage II+ versus benign 0.83, 0.69-0.94; Q P=0.047), reflecting higher discrimination for stage I disease. Heterogeneity flagged for race is attributable to a degenerate small stratum with only 4 non-White cancers and is not interpretable as differential performance.

### Discrimination by time to diagnosis

To assess whether LungAI’s advantage is confined to cancers a radiologist would likely diagnose promptly unaided, we stratified discrimination by the interval from examination to cancer diagnosis (<1 year versus ≥1 year; Supplementary Figures S6–S7; Table S13). At baseline, LungAI retained useful discrimination among cancers diagnosed ≥1 year after the examination (AUROC 0.86, 95% CI 0.78-0.93), where mean diameter was close to chance (AUROC 0.63, 0.49-0.76; ΔAUROC 0.23, P<0.001). The follow-up configuration showed the same direction of effect (ΔAUROC 0.23, P=0.044), although with only 11 cancers in the ≥1-year stratum its estimates are wide (AUROC 0.78, 0.59-0.96). AUPRC is not comparable across strata because prevalence differs and benign controls are shared.

## Discussion

In a held-out NLST partition identical to the published test splits of the Sybil and Ardila et al. papers, LungAI discriminated screen-detected cancers significantly more accurately than Sybil, mean diameter, and estimated volume. Most significantly, LungAI contributed risk information *orthogonal* to the size and growth features on which Lung-RADS and clinical calculators principally rest. The practical expression of that orthogonality is the management redistribution of Figure 4. Integrated into the existing Lung-RADS algorithm without altering its structure, LungAI decreased the proportion of cancers on the slowest surveillance pathway from 9% to 5%, increased the proportion of cancers triaged directly to tissue sampling from 56% to 73%, reduced the rate of benign nodules triaged directly to tissue sampling by 20%, and increased the positivity rate of the Lung-RADS 4B/4X designation from 36% to 48%. An algorithm that merely re-ranked by size could not plausibly achieve both, as diameter’s standalone AUROC (0.78) and the diameter-adjusted likelihood-ratio test make clear.

Our positioning is deliberately conservative: LungAI is intended to augment Lung-RADS rather than replace it. Radiologists first assign a Lung-RADS category and then use the LungAI score to determine whether to up- or down-triage the examination, requiring no new reporting infrastructure. Because the overwhelming majority of screen-detected nodules are benign and reside in lower Lung-RADS categories, a tool that confidently rules out such nodules can reduce interval imaging and invasive workup, while reliable up-triage of the minority of low-category malignancies shortens time to diagnosis. We believe that this is the realistic adoption path for screening CADx technology, and it frames the appropriate comparison set: not AI versus radiologist, but workflow-with versus workflow-without the AI biomarker.

Two findings deserve emphasis beyond the headline discrimination. First, the follow-up configuration – a transparent, three-variable logistic regression using the standalone model’s output, volume doubling time, and nodule tracking status – shows that where growth information exists, it should be used, and that the imaging biomarker remains independently informative after growth is accounted for (χ^2^=79.4). This fusion layer is interpretable, and the proprietary component is confined to the nodule-level score.

Second, our adherence-adjusted analysis quantifies a mechanism usually discussed qualitatively: patients told that their scan looks benign and to “return in 12 months” frequently do not [24–28], and up-triaging elevated-risk Lung-RADS 2 patients to a 6-month pathway both shortens the interval and concretizes follow-up. Accounting for published re-scan adherence rates, we estimate that LungAI would yield 1.6 additional early diagnoses per 1,000 examinations while generating a modest net cost saving, a favorable profile relative to accepted costs of screening-based early cancer detection [29]. The avoided benign biopsies also carry a direct harm benefit independent of cost: at published complication rates [9], roughly one pneumothorax and 0.4 chest-tube placements are averted per 1,000 nodule-positive examinations. A formal cost-effectiveness analysis with staged treatment costs and quality-adjusted outcomes is warranted, and the figures here should be read as bounding estimates under stated assumptions (Table S7).

Future screening will not rest on imaging alone. Blood-based assays – ctDNA, protein panels, methylation signatures – capture molecular shedding, a biological axis orthogonal to both radiologist reads and imaging AI. The natural evolution of lung cancer screening is Lung-RADS alone → Lung-RADS plus imaging AI (this work) → a unified risk model integrating Lung-RADS, imaging AI and circulating biomarkers, each contributing orthogonal signals. The AI-biomarker framing introduced here generalizes directly: each modality earns its place by the information it adds conditional on previous modalities.

### Limitations

First, this is a single-cohort held-out validation; no external cohort is included, and NLST scanners date to 2002–2004. Multi-site external validation on contemporary data is underway at several institutions. Second, although part-solid nodules were eligible, only 7 (0.3%, no cancers) entered the cohort; conclusions apply to solid nodules, and ground-glass disease is out of scope. Third, stage at diagnosis reflects care delivered in NLST, which precedes modern lung cancer screening, not the hypothetical reclassified pathway. Thus, the stage breakdown by reclassification direction (Table S6) is purely descriptive and cannot establish the effect of reclassification on stage at diagnosis.

## Conclusion

LungAI supplies malignancy-risk information orthogonal to the features underlying Lung-RADS. Deployed as a within-category stratifier on the existing management ladder, LungAI reallocates screening patients so that cancers move toward earlier evaluation and benign patients away from invasive workup, all at unchanged biopsy volume and an estimated net cost saving. External validation will determine whether these operating characteristics transport to other settings.

## Data Availability

NLST data are available through the NCI Cancer Data Access System (CDAS). Sybil is publicly available. The LungAI model is proprietary. Analysis code is available upon reasonable request.

## Supplementary materials

Supplementary Tables: S1 Lung-RADS × LungAI stratum cross-tabulation; S2 nested-model and locked fusion coefficients; S3 sampling design and weights; S4 threshold-perturbation robustness; S5 nodule-level discrimination; S6 stage at diagnosis by reclassification direction; S7 cost assumptions; S8 adherence inputs; S9 cohort characteristics; S10 examination-level discrimination, standalone and follow-up configurations; S11 reclassification performance; S12 subgroup discrimination; S13 discrimination by time from examination to diagnosis. Supplementary Figures: S1 STARD flow diagram; S2 subgroup forest plot; S3 Schema B reclassification tree; S4 nodule-level ROC and precision– recall curves; S5 decision-curve analysis; S6 examination-level discrimination by time to diagnosis; S7 nodule-level discrimination by time to diagnosis.

## Notes

### Competing Interest Statement

HS, YK, JG, BM, NC, TS, and YA are equity owners in Oatmeal Health, which develops LungAI; JG (Chief Executive Officer) and YA (Chief Technology Officer) are officers of the company. BM, the radiologist who assigned Lung-RADS categories, serves as a paid advisor to Oatmeal Health. DM is an advisor to Oatmeal Health. LL was previously a contractor for Oatmeal Health.

### Author Declarations

This study only used only available data from the National Lung Screening Trial (NLST). NLST data are available through the NCI Cancer Data Access System (CDAS). This secondary analysis used de-identified NLST data obtained from the NCI Cancer Data Access System under an approved data-use agreement; no investigator had access to identifiable information, and institutional review board review was therefore not required.

