## Supplementary Materials for "LungAI: An AI Biomarker that Refines Lung-RADS v2022 Triage of Screen-Detected Pulmonary Nodules"

All performance figures derive from a retrospective analysis of the NLST held-out test partition. LungAI is an investigational device; a 510(k) premarket notification is pending with the US Food and Drug Administration, and LungAI is not available for sale in the United States.

#### Supplementary Figure S1. STARD flow diagram

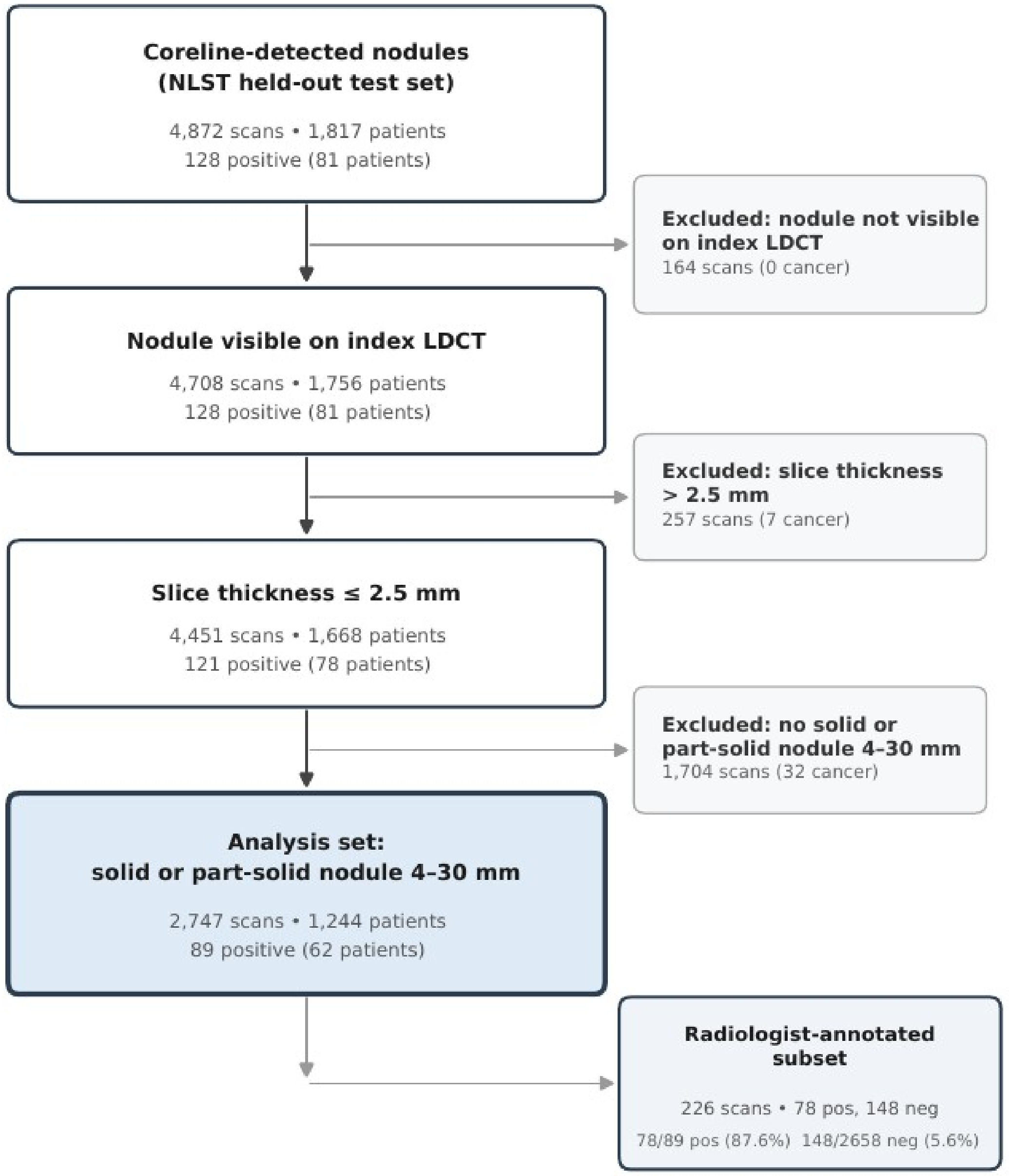


**Figure S1:** *STARD flow diagram. Derivation of the analysis set (2,747 examinations; 89 cancers) from the NLST held-out test partition, and the radiologist-annotated subset (256 annotated; 226 analysis-eligible: 78 cancers, 148 benign) with sampling fractions used for contingency weighting.*

#### Table S1. Lung-RADS × LungAI stratum cross-tabulation

Annotated counts with contingency-weighted projection to the full analysis set; strata are defined by displayed LungAI score cutpoints (Methods).

| Lung-RADS | LungAI stratum | Weighted n (malignant) | Malignant % | Management |
|---|---|---|---|---|
| 2 | <2.0 | 1,331 (2) | 0.2% | 12-mo LDCT |
| 2 | ≥2.0 | 473 (7) | 1.3% | 6-mo LDCT |
| 3 | <1.0 | 254 (2) | 0.9% | 12-mo LDCT |
| 3 | 1.0–3.9 | 200 (2) | 1.2% | 6-mo LDCT |
| 3 | ≥4.0 | 22 (4) | 16.7% | 3-mo LDCT/PET |
| 4A | <1.5 | 109 (1) | 1.0% | 6-mo LDCT |
| 4A | 1.5–5.9 | 185 (6) | 3.0% | 3-mo LDCT/PET |
| 4A | ≥6.0 | 33 (15) | 46.2% | Biopsy/resection |
| 4B/4X | <2.0 | 36 (0) | 0.0% | 3-mo LDCT/PET |
| 4B/4X | ≥2.0 | 104 (50) | 48.3% | Biopsy/resection |

#### Table S2. Model Coefficients

##### S2a. Nested orthogonality models

**Baseline** (n=2,747; 89 events): adding the LungAI score to a restricted-cubic-spline model of diameter yields likelihood-ratio $\chi^2$ = 161.0, P<0.001; ΔAUROC = 0.104 (95% CI 0.057–0.179). Diameter spline terms remain significant in the joint model, indicating residual size information independent of LungAI.

| Variable | Coefficient | OR (95% CI) | P |
|---|---|---|---|
| Intercept | −3.525 | 0.029 (0.018–0.047) | <0.001 |
| $rcs(diameter)_1$ | −1.579 | 0.206 (0.136–0.314) | <0.001 |
| $rcs(diameter)_2$ | −0.930 | 0.394 (0.266–0.585) | <0.001 |
| $rcs(diameter)_3$ | −1.017 | 0.362 (0.133–0.986) | 0.047 |
| LungAI score | 7.821 | 2,492 (592–10,484) | <0.001 |

**Follow-up** (n=1,899; 45 events): adding the LungAI baseline score to M0 = rcs(diameter) + volume doubling time + new-nodule indicator yields likelihood-ratio $\chi^2$ = 79.4, P<0.001; ΔAUROC = 0.060 (95% CI 0.023–0.131). In the joint model, the diameter spline terms and volume doubling time (OR 2.54 per doubling per year, 95% CI 1.69–3.81; P<0.001) remain independently significant, while the new-nodule indicator (OR 0.51, 95% CI 0.22–1.17;

P=0.113) does not. This suggests that growth dynamics contribute information beyond what LungAI captures from a single examination.

| Variable | Coefficient | OR (95% CI) | P |
|---|---|---|---|
| Intercept | −3.935 | 0.020 (0.009–0.044) | <0.001 |
| $rcs(diameter)_1$ | −1.352 | 0.259 (0.133–0.504) | <0.001 |
| $rcs(diameter)_2$ | −0.742 | 0.476 (0.248–0.915) | 0.026 |
| $rcs(diameter)_3$ | −1.841 | 0.159 (0.029–0.872) | 0.034 |
| Volume doubling time (doublings per year) | 0.931 | 2.536 (1.691–3.805) | <0.001 |
| New nodule (vs tracked) | −0.674 | 0.510 (0.221–1.173) | 0.113 |
| LungAI score | 8.726 | 6,160 (662–57,335) | <0.001 |

##### S2b. Locked follow-up fusion coefficients (configuration C1)

$z = \beta_0 + \beta_1 \cdot \text{logit(baseline probability)} + \beta_2 \cdot \text{(volume doubling time)} \cdot I(\text{tracked}) + \beta_3 \cdot I(\text{tracked})$; follow-up probability = sigmoid(0.9556·z + 0.3803)

| Variable | Locked coefficient | OR |
|---|---|---|
| Intercept | −1.0171 | 0.362 |
| logit(baseline probability) | 0.8828 | 2.418 |
| Volume doubling time × tracked, per doubling per year | 0.9665 | 2.629 |
| Tracked indicator | 0.2199 | 1.246 |

Coefficients were fit and cross-validated on the NLST validation partition and locked before any evaluation of the test partition; locked values were recovered exactly from device outputs ($R^2 = 1.000$). Volume doubling time is $\log_2$(Vcurrent/Vprior) per year with the inter-examination interval fixed at $\Delta t = 1$ year (observed NLST inter-scan interval median 1.00 year, IQR 0.94–1.06); untracked nodules carry no growth term, and tracked nodules whose prior detection has no measurement receive the tracking indicator with the growth term set to zero (458 nodules, all benign).

### Supplementary Figure S2. Subgroup forest plot

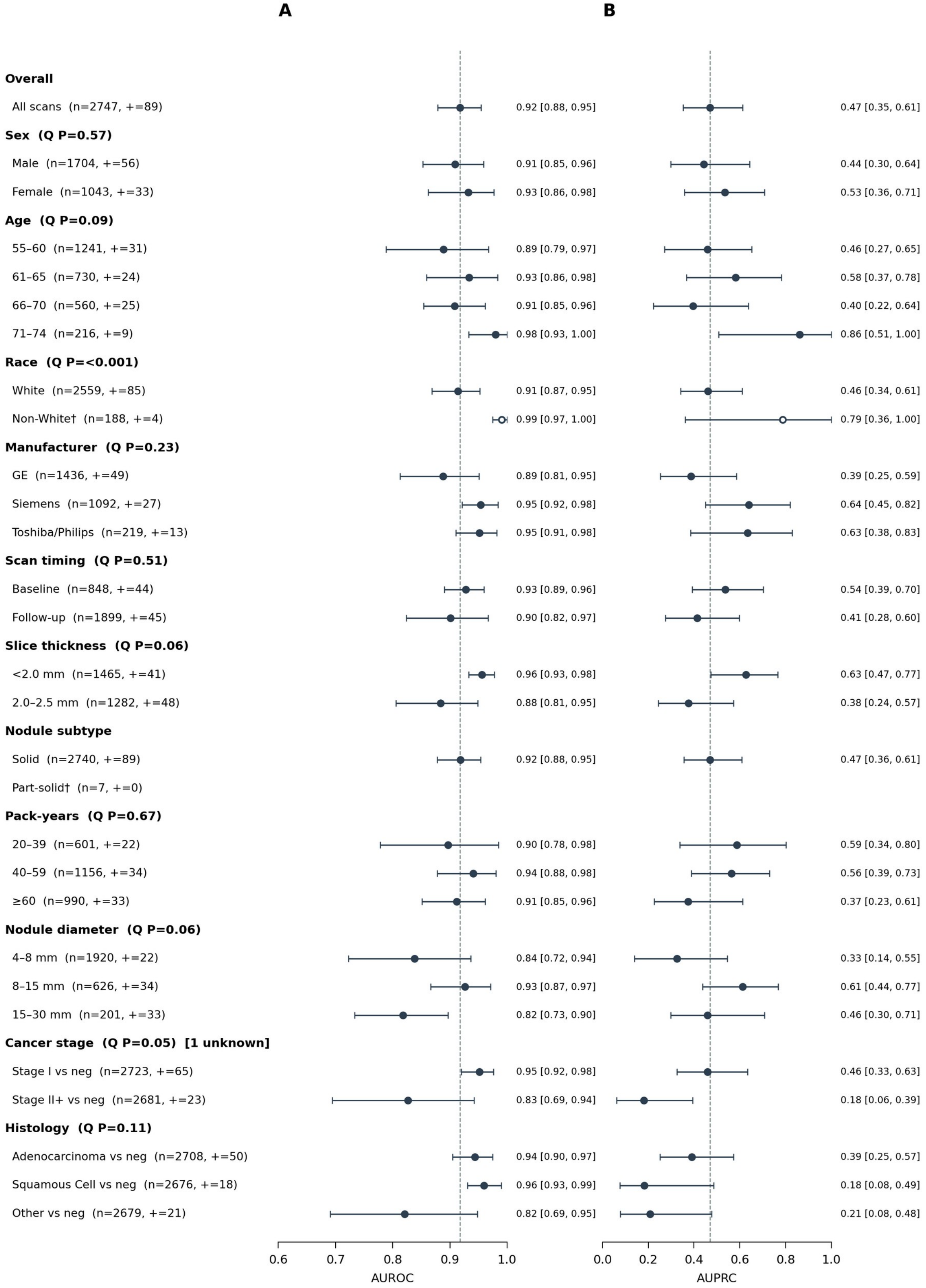

**Figure S2:** *Subgroup forest plot. Examination-level (A) AUROC and (B) AUPRC (95% CI) with Cochran's Q per subgroup. † denotes degenerate small strata.*

#### Table S3. Annotated-subset sampling design and contingency weights

| Quantity | Value |
|---|---|
| Analysis set | 2,747 examinations (89 malignant, 2,658 benign) |
| Annotated by radiologist | 256 examinations |
| Annotated and in analysis set | 226 examinations (78 malignant, 148 benign) |
| Annotated but analysis-ineligible (excluded) | 30 examinations |
| Sampling fraction, malignant | 78/89 = 87.6% |
| Sampling fraction, benign | 148/2,658 = 5.6% |
| Contingency weight, malignant ($w^+$ = 89/78) | 1.14 |
| Contingency weight, benign ($w^-$ = 2,658/148) | 17.96 |

All Lung-RADS–dependent quantities (within-category performance, cross-tabulations, reclassification counts) are projected to the full analysis set by applying $w^+/w^-$ to annotated examinations; confidence intervals by weighted patient-clustered bootstrap.

#### Supplementary Figure S3. Schema B reclassification threshold tree

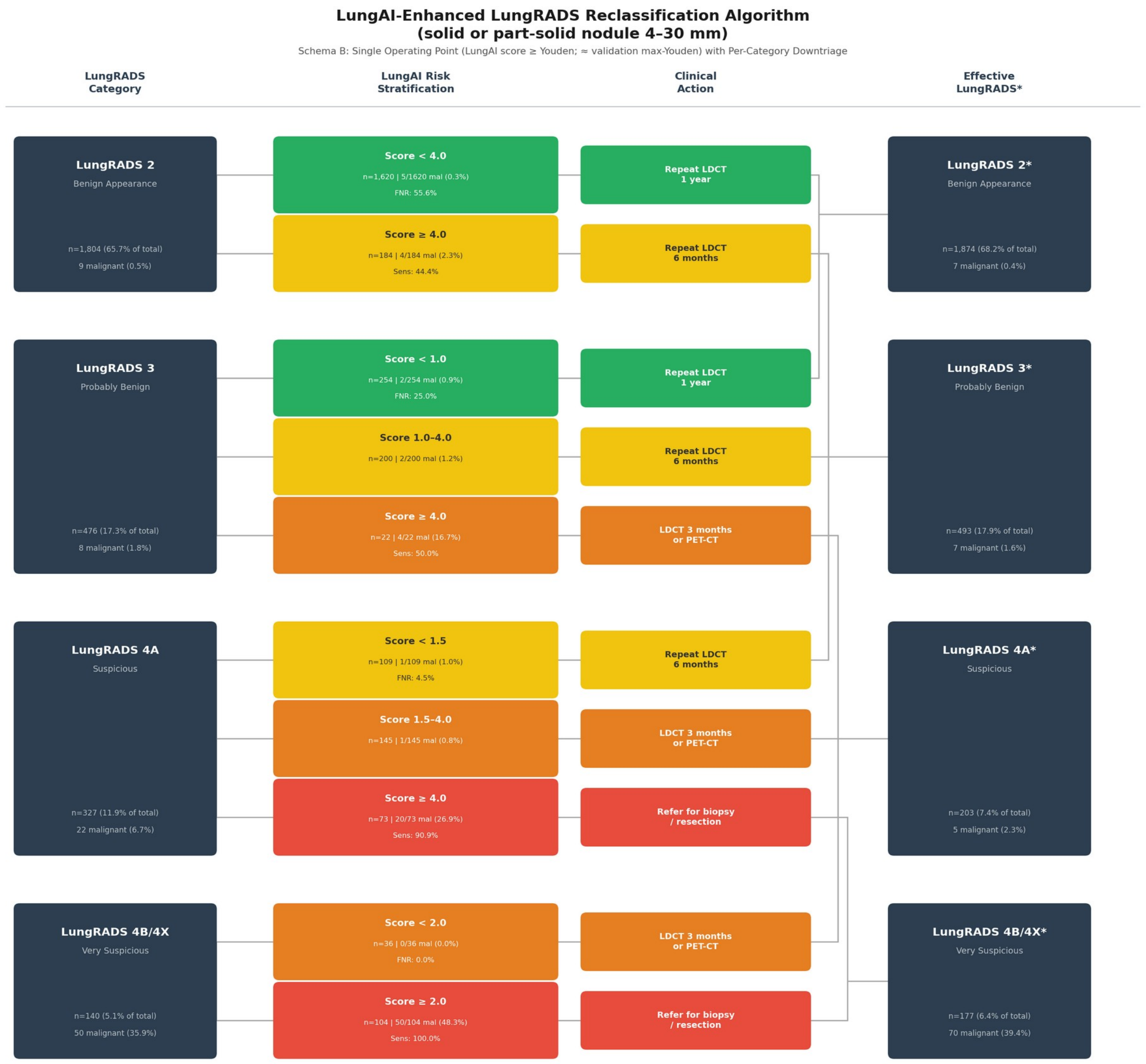


**Figure S3:** *Schema B: Reclassification within Lung-RADS categories using a single up-triage operating point at LungAI score 4.0 across all categories (approximating the maximum-Youden threshold derived on the NLST validation partition), with per-category down-triage cutpoints unchanged. Layout as in main-text Figure 3.*

#### Table S4. Threshold-perturbation robustness (Schema A, joint scaling of all six decision thresholds)

Per 1,000 nodule-positive examinations. The operating thresholds of all six cutpoints were scaled by factors from 0.50 to 2.00.

| Threshold multiplier | Cancers on 12-mo pathway | Benign biopsies | Total biopsies | Net lead time (mo) | Event NRI | Non-event NRI |
|---|---|---|---|---|---|---|
| ×0.50 | 1.6 | 39.2 | 63.5 | 27.5 | +0.270 | −0.142 |
| ×0.75 | 1.6 | 32.7 | 57.0 | 27.5 | +0.270 | −0.068 |
| **×1.00 (primary)** | **1.6** | **26.2** | **50.1** | **25.2** | **+0.245** | **−0.041** |
| ×1.25 | 1.6 | 26.2 | 48.8 | 22.2 | +0.208 | −0.007 |
| ×1.50 | 1.6 | 13.1 | 35.4 | 21.2 | +0.196 | +0.027 |
| ×2.00 | 1.6 | 13.1 | 34.2 | 18.3 | +0.159 | +0.074 |

Event NRI remains positive (>0.15) across the entire four-fold range. Cancers on the 12-month pathway remain at 1.6 versus 3.1 under Lung-RADS alone; net lead time remains 18-28 months. At the most conservative thresholds (×1.50 and above), the non-event NRI itself turns positive. This suggests a significantly positive NRI robust to threshold perturbation.

#### Table S5. Nodule-level discrimination

For our nodule-level analysis, each cancer examination contributes one nodule – the diagnosed target nodule – while benign examinations contribute all nodules meeting inclusion criteria. Since including multiple nodules per examination violates the independence assumption of the DeLong test, all nodule-level inference uses patient-clustered bootstrap (2,000 replicates) for both confidence intervals and P values. Sybil produces no nodule-level output so is omitted.

**Table S5a: Discrimination, standalone configuration.**

| **Model** | **AUROC (95% CI)** | **AUPRC (95% CI)** | **ΔAUROC (95% CI); P-value** | **ΔAUPRC (95% CI); P-value** |
|---|---|---|---|---|
| LungAI | 0.93 (0.89-0.96) | 0.37 (0.23-0.57) | ref | ref |
| Mean diameter | 0.84 (0.78-0.90) | 0.11 (0.07-0.19) | 0.09 (0.03-0.15); P=0.001 | 0.26 (0.12-0.44); P<0.001 |
| Volume | 0.84 (0.77-0.90) | 0.10 (0.07-0.16) | 0.10 (0.04-0.17); P=0.001 | 0.29 (0.15-0.47); P<0.001 |

**Table S5b: Discrimination, follow-up configuration (4,242 nodules, 45 malignant).**

| **Model** | **AUROC (95% CI)** | **AUPRC (95% CI)** | **ΔAUROC (95% CI); P** | **ΔAUPRC (95% CI); P** |
|---|---|---|---|---|
| LungAI-FU | 0.91 (0.84-0.98) | 0.51 (0.34-0.72) | ref | ref |

| | | | | |
|---|---|---|---|---|
| LungAI (standalone) | 0.91 (0.84-0.97) | 0.30 (0.17-0.55) | 0.01 (0.00-0.01); P=0.049 | 0.21 (0.10-0.28); P<0.001 |
| Δ diameter‡ | 0.88 (0.78-0.97) | 0.37 (0.20-0.57) | 0.09 (0.03-0.17); P<0.001 | 0.38 (0.17-0.56); P=0.001 |
| Δ volume‡ | 0.89 (0.78-0.97) | 0.39 (0.21-0.61) | 0.08 (0.02-0.17); P<0.001 | 0.36 (0.16-0.50); P=0.001 |

‡ Δdiameter and Δvolume are defined only for tracked nodules (matched to a prior detection) and are evaluated on that subset (1,664 nodules, 27 malignant). Paired deltas compare LungAI-FU with each comparator on the same nodules.

Nodule-level AUROC exceeds examination-level AUROC (0.93 vs 0.92) because the per-nodule score classifies the target nodule directly, without max-aggregation over benign nodules in multi-nodule examinations. Nodule-level AUPRC is lower (0.37 vs 0.47) because nodule-level prevalence is lower (1.5% vs 3.2%). The precision advantage of the follow-up configuration replicates at nodule level (ΔAUPRC +0.21 versus the standalone score on identical nodules, P<0.001), as does the pattern against growth metrics: LungAI-FU significantly exceeds both Δdiameter and Δvolume on AUROC and AUPRC on the tracked-nodule subset (all P≤0.001). Nodule-level ROC and PRC curves are in Supplementary Figure S4.

#### Supplementary Figure S4. Nodule-level discrimination curves

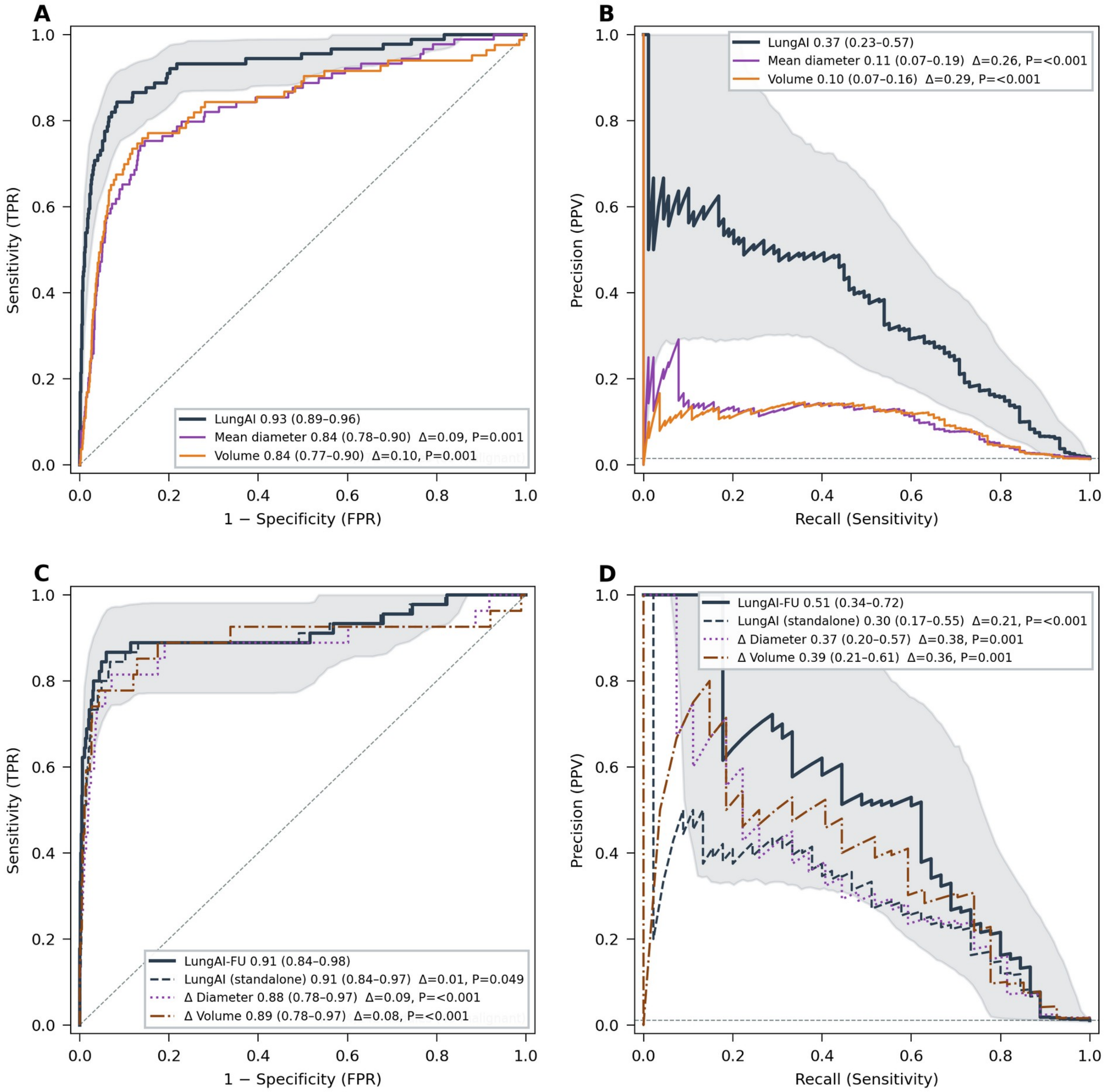


***Figure S4:*** *Nodule-level ROC (A, C) and precision-recall (B, D) curves for the baseline (A-B; 5,876 nodules, 89 malignant) and follow-up (C-D; 4,242 nodules, 45 malignant) configurations versus size- and growth-based comparators; patient-clustered bootstrap confidence intervals.*

#### Table S6. Stage at diagnosis by reclassification direction

Stage at diagnosis by direction of LungAI-based reclassification among radiologist-annotated cancers (Schema A). Stage is the AJCC 7th-edition stage from NLST assigned per patient to each cancer examination. Of 78 annotated cancers, 20 were up-triaged to a more intensive management category, 55 were unchanged, and 3 were down-triaged. The two narrow rows isolate cancers moved off the 12-month pathway (Lung-RADS 2 with LungAI score ≥2.0) and cancers remaining on it (Lung-RADS 2 with score <2.0, or Lung-RADS 3 down-triaged to 12 months). Counts are unweighted annotated cancers with contingency-weighted totals alongside.

| **Group** | **Annotated n** | **Weighted n** | **Stage I** | **IA / IB** | **Stage II+** | **Unknown** | **% stage I of staged** |
|---|---|---|---|---|---|---|---|
| All annotated cancers | 78 | 89.0 | 58 | 41 / 17 | 19 | 1 | 75.3% |
| Up-triaged | 20 | 25.3 | 16 | 10 / 6 | 4 | 0 | 80.0% |
| Unchanged | 55 | 60.2 | 41 | 31 / 10 | 13 | 1 | 75.9% |
| Down-triaged | 3 | 3.5 | 1 | 0 / 1 | 2 | 0 | 33.3% |
| Up-triaged off 12-month pathway (LR 2, score ≥2.0) | 3 | 6.3 | 2 | 2 / 0 | 1 | 0 | 66.7% |
| Remaining on 12-month pathway | 2 | 2.4 | 1 | 0 / 1 | 1 | 0 | 50.0% |

Exploratory Fisher exact test (up-triaged versus all others, stage II+ versus stage I): 4/20 versus 15/57, OR 0.70, P=0.77 (unadjusted for patient clustering). Stage reflects care delivered in NLST and tumor biology, not the hypothetical reclassified pathway, and the LungAI score is itself associated with tumor aggressiveness. These distributions are descriptive and cannot establish the effect of reclassification on stage at diagnosis.

#### Supplementary Figure S5. Decision-curve analysis

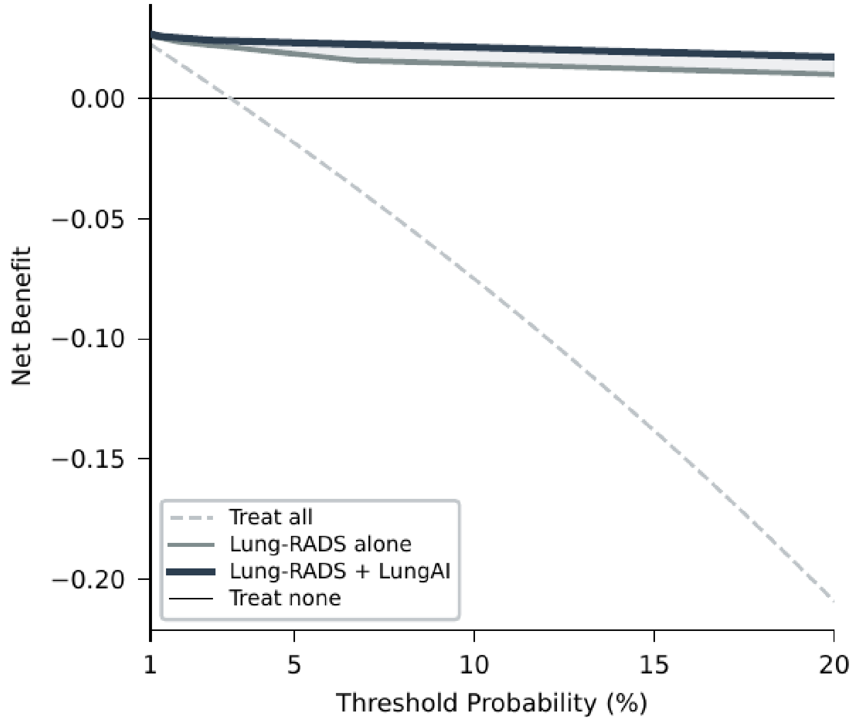


**Figure S5:** *Net benefit versus threshold probability (1-20%) for the combined Lung-RADS + LungAI algorithm versus Lung-RADS alone, treat-all, and treat-none. Probability inputs are stratum-level observed malignant fractions from the weighted cross-tabulation; because Lung-RADS categories exist only in the annotated evaluation subset, these risk estimates are in-sample, and the comparison is descriptive rather than out-of-sample.*

#### Table S7. Cost assumptions

| Item | Value | Basis |
|---|---|---|
| LDCT examination | $300 | Medicare, CPT 71271 |
| CT-guided biopsy | $4,000 | Midpoint of $3,000 – $5,000 range, consistent with a Medicare claims analysis reporting a median of ~$3,800 per biopsy (Lokhandwala T, et al. Clin Lung Cancer. 2017; 18(1): e27-e34) |
| 12-month management | $0 incremental | Annual screening occurs regardless |
| 6-month / 3-month management | +$300 | One additional interval LDCT |
| Biopsy cost attribution | Benign patients only | Cancer patients undergo biopsy at eventual diagnosis regardless |

#### Table S8. Follow-up adherence inputs

"Defined adherence" (completion within the recommended window plus allowed latitude), unweighted mean ± SD across four studies identified in the meta-analysis of Lin et al. (J Thorac Oncol 2022): Barbosa (AJR 2021), Bernstein (Am J Respir Crit Care Med 2019, abstract), Lake (J Am Coll Radiol 2020), Triplette (Ann Am Thorac Soc 2021).

| Lung-RADS | Management | Defined adherence |
|---|---|---|
| 2 | 12-month LDCT | 37.9% ± 21.7% |
| 3 | 6-month LDCT | 64.1% ± 9.0% |
| 4A | 3-month LDCT / PET-CT | 81.3% ± 8.7% |
| 4B/4X | Biopsy referral | 90.1% ± 8.4% |

Structural assumption: patients on a 12-month re-scan pathway have, by definition, no opportunity for diagnosis within 1 year.

#### Table S9. Cohort characteristics (NLST held-out analysis set)

| Characteristic | Full (n=2,747) | Cancer (n=89) | Non-cancer (n=2,658) |
|---|---|---|---|
| Age, median (IQR), y | 61 (58-66) | 63 (59-67) | 61 (57-66) |
| Sex: male | 1,704 (62.0%) | 56 (62.9%) | 1,648 (62.0%) |
| Sex: female | 1,043 (38.0%) | 33 (37.1%) | 1,010 (38.0%) |
| Race: White | 2,559 (93.2%) | 85 (95.5%) | 2,474 (93.1%) |
| Race: Black or African-American | 75 (2.7%) | 2 (2.2%) | 73 (2.7%) |
| Race: Asian | 32 (1.2%) | 1 (1.1%) | 31 (1.2%) |
| Race: Other | 66 (2.4%) | 1 (1.1%) | 65 (2.5%) |
| Race: Unknown or not reported | 15 (0.5%) | 0 (0.0%) | 15 (0.6%) |
| Ethnicity: Hispanic or Latino | 41 (1.5%) | 1 (1.1%) | 40 (1.5%) |
| Pack-years, median (IQR) | 50 (40-68) | 50 (41-81) | 49 (40-68) |
| Smoking: current | 1,384 (50.4%) | 45 (50.6%) | 1,339 (50.4%) |
| Smoking: former | 1,363 (49.6%) | 44 (49.4%) | 1,319 (49.6%) |
| Manufacturer: GE | 1,436 (52.3%) | 49 (55.1%) | 1,387 (52.2%) |
| Manufacturer: Siemens | 1,092 (39.8%) | 27 (30.3%) | 1,065 (40.1%) |
| Manufacturer: Toshiba/Philips | 219 (8.0%) | 13 (14.6%) | 206 (7.8%) |

| Characteristic | Full (n=2,747) | Cancer (n=89) | Non-cancer (n=2,658) |
|---|---|---|---|
| Subtype: solid | 2,740 (99.7%) | 89 (100.0%) | 2,651 (99.7%) |
| Subtype: part-solid | 7 (0.3%) | 0 (0.0%) | 7 (0.3%) |

#### Table S10. Examination-level discrimination, standalone and follow-up configurations

**Table S10a. Discrimination, standalone configuration.** ΔAUROC and DeLong P computed on each comparator's common-support (paired) subset; ΔAUPRC differences by paired patient-clustered bootstrap (2,000 replicates). Lung-RADS v2022 scored ordinally on the 226-examination annotated subset with contingency weighting; both AUROC and AUPRC P-values computed by patient-clustered bootstrap.

| Model | AUROC (95% CI) | AUPRC (95% CI) | ΔAUROC (95% CI) | P | ΔAUPRC (95% CI) | P |
|---|---|---|---|---|---|---|
| LungAI | 0.92 (0.88-0.95) | 0.47 (0.35-0.61) | ref | — | ref | — |
| Sybil 1-yr | 0.85 (0.79-0.90) | 0.25 (0.17-0.39) | 0.07 (0.04-0.09) | <0.001 | 0.18 (0.11-0.25) | <0.001 |
| Mean diameter | 0.78 (0.72-0.85) | 0.13 (0.09-0.21) | 0.14 (0.08-0.19) | <0.001 | 0.34 (0.22-0.47) | <0.001 |
| Volume | 0.79 (0.72-0.86) | 0.12 (0.08-0.18) | 0.14 (0.09-0.20) | <0.001 | 0.38 (0.26-0.50) | <0.001 |
| Lung-RADS v2022† | 0.89 (0.82-0.94) | 0.25 (0.15-0.47) | 0.03 (-0.04-0.10) | 0.20 | 0.31 (0.10-0.50) | 0.003 |

† Lung-RADS v2022 scored ordinally on the 226-examination annotated subset with contingency weighting; ΔAUROC and ΔAUPRC P-values by patient-clustered bootstrap.

**Table S10b. Discrimination, follow-up configuration.** ΔAUROC and DeLong P computed on each comparator's common-support (paired) subset on follow-up scans only (n=1,899; 45 cancers); ΔAUPRC differences by paired patient-clustered bootstrap (2,000 replicates).

| Model | AUROC (95% CI) | AUPRC (95% CI) | ΔAUROC (95% CI) | P | ΔAUPRC (95% CI) | P |
|---|---|---|---|---|---|---|
| LungAI-FU | 0.91 (0.83-0.97) | 0.60 (0.45-0.74) | ref | — | ref | — |
| LungAI (on follow-up scans) | 0.90 (0.82-0.97) | 0.41 (0.28-0.60) | 0.01 (0.00-0.02) | 0.13 | 0.18 (0.09-0.26) | <0.001 |

| Model | AUROC (95% CI) | AUPRC (95% CI) | ΔAUROC (95% CI) | P | ΔAUPRC (95% CI) | P |
|---|---|---|---|---|---|---|
| Sybil 1-yr | 0.84 (0.74-0.92) | 0.19 (0.11-0.35) | 0.06 (0.03-0.09) | <0.001 | 0.37 (0.24-0.46) | <0.001 |
| Δ diameter‡ | 0.86 (0.76-0.95) | 0.37 (0.20-0.57) | 0.11 (0.03-0.19) | 0.009 | 0.36 (0.16-0.53) | 0.001 |
| Δ volume‡ | 0.88 (0.77-0.96) | 0.40 (0.22-0.62) | 0.09 (0.01-0.17) | 0.021 | 0.32 (0.13-0.48) | 0.001 |

‡ Δdiameter and Δvolume are defined only for tracked nodules with a measured prior and are evaluated on the 963 follow-up examinations with at least one such nodule (27 cancers). The paired ΔAUROC and ΔAUPRC compare LungAI-FU with each comparator on these same examinations, while LungAI-FU's own AUROC and AUPRC are reported on the full follow-up set (n=1,899).

#### Table S11. Reclassification performance of the combined algorithm versus Lung-RADS v2022 alone

McNemar's exact test on the annotated subset. Net reclassification index (NRI) weighted with patient-clustered bootstrap CIs.

| Measure | Estimate | 95% CI / P |
|---|---|---|
| McNemar, cancers: up-triaged / down-triaged | 20 / 3 | P<0.001 |
| McNemar, benign: up-triaged / down-triaged | 28 / 22 | P=0.480 |
| Event NRI | +0.25 | 0.13-0.37 |
| Non-event NRI | -0.04 | -0.13 to 0.05 |
| Overall NRI | +0.20 | 0.04-0.36 |
| Cancers on 12-month rescan pathway (contingency weighted), before → after | 8.4 → 4.5 | — |
| Benign biopsy referrals (contingency weighted), before → after | 90 → 72 | — |

#### Table S12. Subgroup discrimination (baseline configuration)

Examination-level AUROC (95% CI); heterogeneity by Cochran's Q with $I^2$. † degenerate small stratum (n<30 or <5 events). Stage comparisons exclude 1 cancer with unknown stage.

| Stratum | n (pos) | AUROC (95% CI) |
|---|---|---|
| All examinations | 2,747 (89) | 0.92 (0.88-0.95) |

| Stratum | n (pos) | AUROC (95% CI) |
|---|---|---|
| **Sex** (Q=0.33, P=0.57, $I^2$=0%) | | |
| Male | 1,704 (56) | 0.91 (0.85-0.96) |
| Female | 1,043 (33) | 0.93 (0.86-0.98) |
| **Age** (Q=6.47, P=0.091, $I^2$=53.7%) | | |
| 55-60 | 1,241 (31) | 0.89 (0.79-0.97) |
| 61-65 | 730 (24) | 0.93 (0.86-0.98) |
| 66-70 | 560 (25) | 0.91 (0.85-0.96) |
| 71-74 | 216 (9) | 0.98 (0.93-1.00) |
| **Scanner manufacturer** (Q=2.91, P=0.23, $I^2$=31.3%) | | |
| GE | 1,436 (49) | 0.89 (0.81-0.95) |
| Siemens | 1,092 (27) | 0.95 (0.92-0.98) |
| Toshiba/Philips | 219 (13) | 0.95 (0.91-0.98) |
| **Scan timing** (Q=0.44, P=0.51, $I^2$=0%) | | |
| Baseline round | 848 (44) | 0.93 (0.89-0.96) |
| Follow-up rounds | 1,899 (45) | 0.90 (0.82-0.97) |
| **Slice thickness (Q=3.59, P=0.058, $I^2$=72.1%)** | | |
| <2.0 mm | 1,465 (41) | 0.96 (0.93-0.98) |
| 2.0-2.5 mm | 1,282 (48) | 0.88 (0.80-0.95) |
| **Race (Q=11.43, P<0.001, $I^2$=91.3%)†** | | |
| White | 2,559 (85) | 0.91 (0.87-0.95) |
| Non-White† | 188 (4) | 0.99 (0.97-1.00) |
| **Nodule subtype** | | |
| Solid | 2,740 (89) | 0.92 (0.88-0.96) |
| Part-solid† | 7 (0) | not computable |
| **Pack-years** (Q=0.81, P=0.67, $I^2$=0%) | | |
| 20-39 | 601 (22) | 0.90 (0.78-0.98) |

| Stratum | n (pos) | AUROC (95% CI) |
|---|---|---|
| 40-59 | 1,156 (34) | 0.94 (0.88-0.98) |
| ≥60 | 990 (33) | 0.91 (0.85-0.96) |
| **Nodule diameter** (Q=5.52, P=0.063, $I^2$=63.8%) | | |
| 4-8 mm | 1,920 (22) | 0.84 (0.72-0.94) |
| 8-15 mm | 626 (34) | 0.93 (0.86-0.97) |
| 15-30 mm | 201 (33) | 0.82 (0.73-0.89) |
| **Cancer stage (Q=3.95, P=0.047, $I^2$=74.7%)** | | |
| Stage I vs benign | 2,723 (65) | 0.95 (0.92-0.98) |
| Stage II+ vs benign | 2,681 (23) | 0.83 (0.69-0.94) |
| **Histology (Q=4.50, P=0.11, $I^2$=55.5%)** | | |
| Adenocarcinoma vs benign | 2,708 (50) | 0.94 (0.90-0.97) |
| Squamous cell vs benign | 2,676 (18) | 0.96 (0.93-0.99) |
| Other vs benign | 2,679 (21) | 0.82 (0.70-0.95) |

#### Supplementary Figures S6–S7 and Table S13. Discrimination by time from examination to diagnosis

To assess whether LungAI's value is driven by examinations a radiologist would likely diagnose promptly anyway, we stratified discrimination by the interval from the examination to cancer diagnosis (diagnosis date minus examination date from the NLST person file): cancers diagnosed <1 year versus ≥1 year after the examination. Benign examinations appear in both strata. Figure S6 reports examination-level and Figure S7 nodule-level results; each shows the baseline and follow-up configurations with ROC and precision-recall curves against mean diameter. Confidence intervals and P values use patient-clustered bootstrap (2,000 replicates). AUPRC is not comparable across strata because prevalence differs and benign controls are shared; AUROC is the comparable metric.

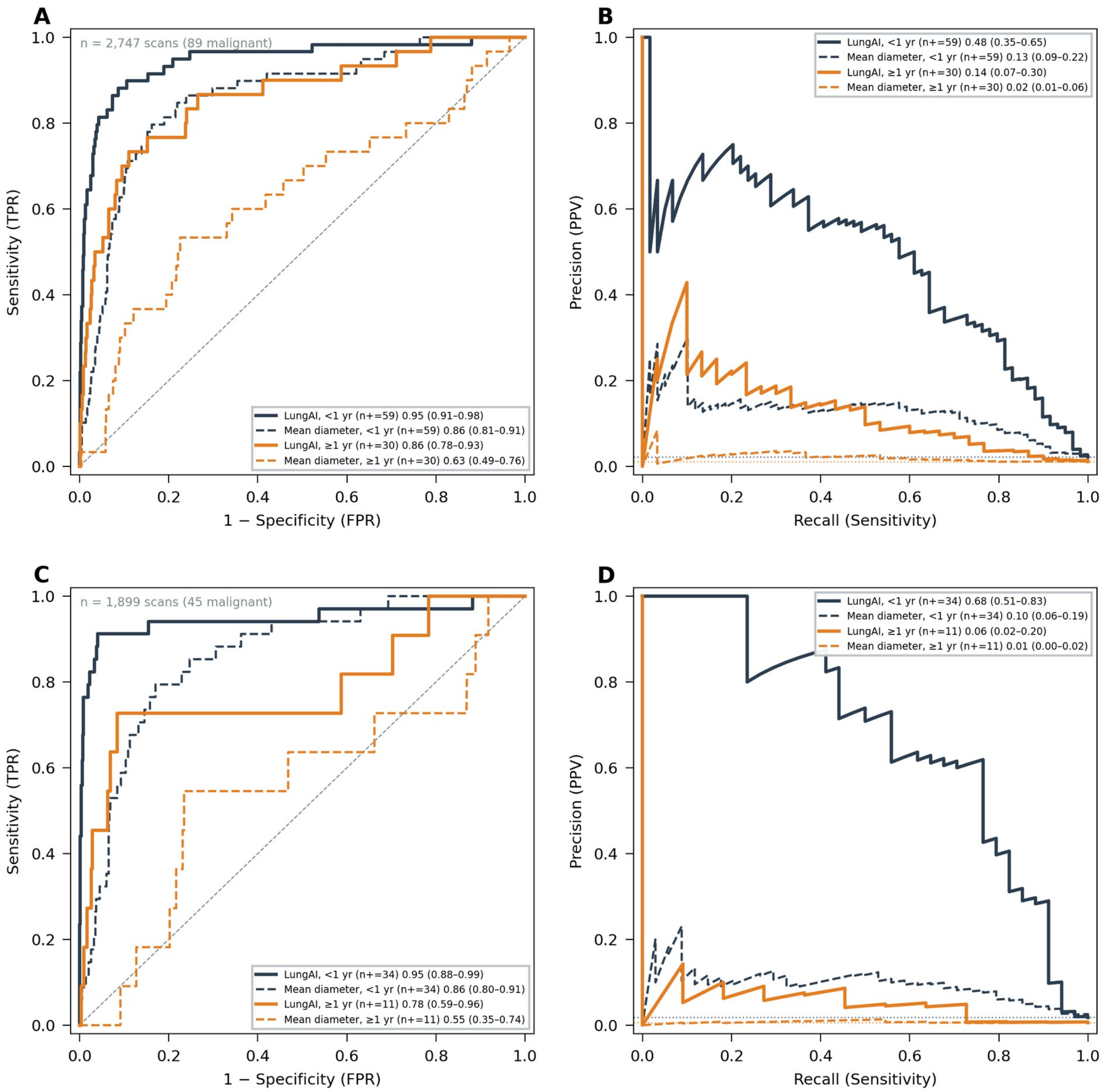


***Figure S6:*** *Examination-level discrimination by time from examination to diagnosis. ROC and precision-recall curves for LungAI versus mean diameter among cancers diagnosed <1 year and ≥1 year after the examination, for the baseline configuration (A-B, 2,747 examinations, 89 cancers: 59 diagnosed <1 year, 30 ≥1 year) and the follow-up configuration (C-D, 1,899 examinations, 45 cancers: 34 diagnosed <1 year, 11 ≥1 year).*

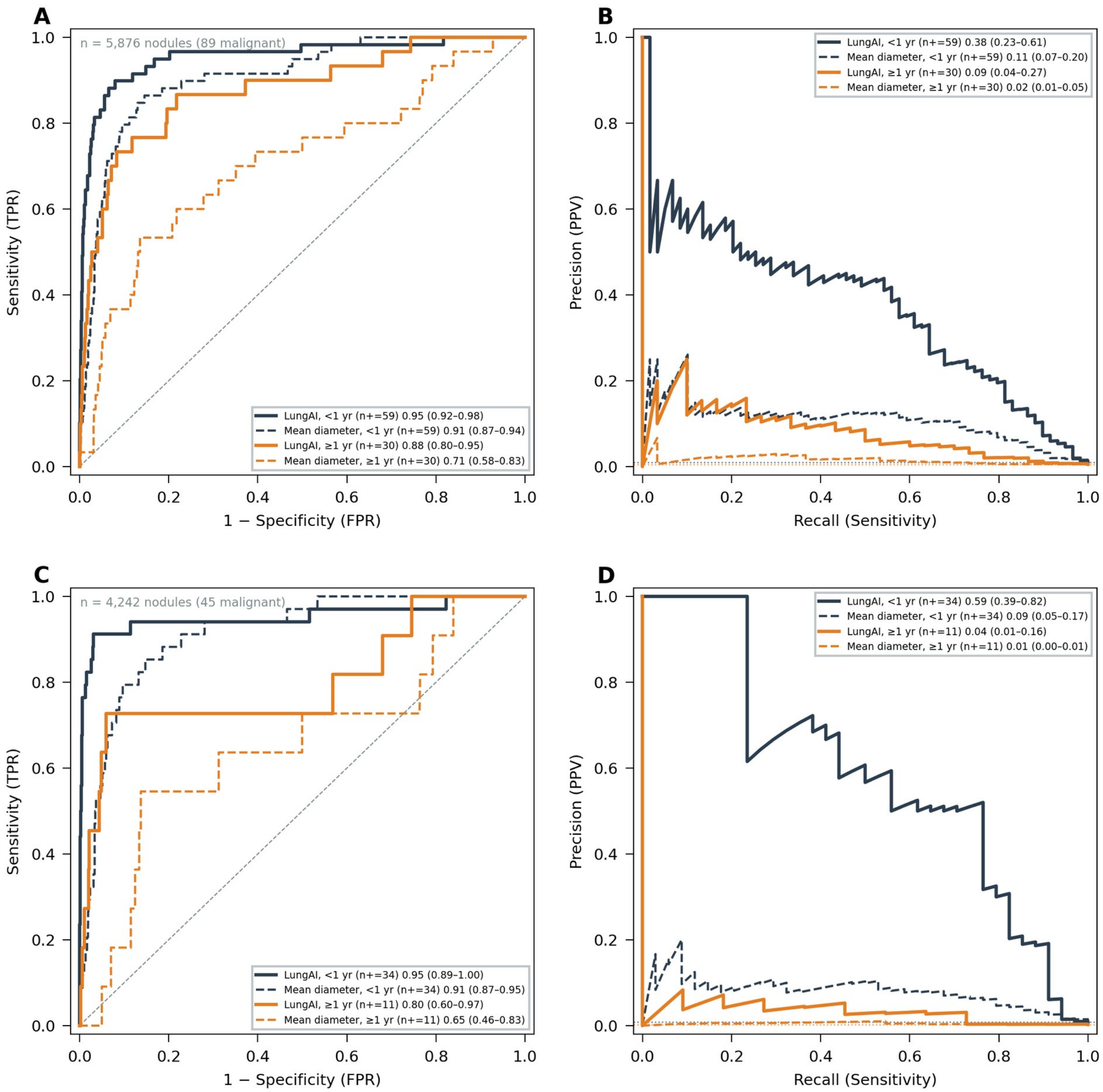


***Figure S7:*** *Nodule-level discrimination by time from examination to diagnosis. Layout as in Figure S6, at nodule level for the standalone (A-B, 5,876 nodules, 89 malignant) and follow-up (C-D, 4,242 nodules, 45 malignant) configurations.*

**Table S13.** Discrimination by time from examination to diagnosis. Patient-clustered bootstrap (2,000 replicates) with mean diameter as comparator.

| Level | Configuration | Stratum | n | Cancers | LungAI AUROC (95% CI) | LungAI AUPRC (95% CI) | Diameter AUROC (95% CI) | ΔAUROC (95% CI) | P |
|---|---|---|---|---|---|---|---|---|---|
| Examination | Baseline | All | 2,747 | 89 | 0.92 (0.88-0.95) | 0.47 (0.35-0.61) | 0.78 (0.72-0.85) | 0.14 (0.07-0.20) | <0.001 |
| Examination | Baseline | <1 yr | 2,717 | 59 | 0.95 (0.91-0.98) | 0.48 (0.35-0.65) | 0.86 (0.81-0.91) | 0.09 (0.04-0.14) | <0.001 |
| Examination | Baseline | ≥1 yr | 2,688 | 30 | 0.86 (0.78-0.93) | 0.14 (0.07-0.30) | 0.63 (0.49-0.76) | 0.23 (0.11-0.37) | <0.001 |
| Examination | Follow-up | All | 1,899 | 45 | 0.91 (0.83-0.97) | 0.60 (0.45-0.74) | 0.79 (0.71-0.86) | 0.12 (0.04-0.20) | 0.001 |
| Examination | Follow-up | <1 yr | 1,888 | 34 | 0.95 (0.88-0.99) | 0.68 (0.51-0.83) | 0.86 (0.80-0.91) | 0.09 (0.04-0.14) | <0.001 |
| Examination | Follow-up | ≥1 yr | 1,865 | 11 | 0.78 (0.59-0.96) | 0.06 (0.02-0.20) | 0.55 (0.35-0.74) | 0.23 (0.01-0.46) | 0.044 |
| Nodule | Baseline | <1 yr | 5,846 | 59 | 0.95 (0.92-0.98) | 0.38 (0.23-0.61) | 0.91 (0.87-0.94) | 0.05 (0.01-0.09) | 0.006 |
| Nodule | Baseline | ≥1 yr | 5,817 | 30 | 0.88 (0.80-0.95) | 0.09 (0.04-0.27) | 0.71 (0.58-0.83) | 0.17 (0.05-0.30) | 0.001 |
| Nodule | Follow-up | <1 yr | 4,231 | 34 | 0.95 (0.89-1.00) | 0.59 (0.39-0.82) | 0.91 (0.87-0.95) | 0.04 (-0.01-0.09) | 0.074 |
| Nodule | Follow-up | ≥1 yr | 4,208 | 11 | 0.80 (0.60-0.97) | 0.04 (0.01-0.16) | 0.65 (0.46-0.83) | 0.15 (-0.07-0.38) | 0.193 |

At baseline, LungAI retained useful discrimination among cancers diagnosed ≥1 year after the examination (examination-level AUROC 0.86, 95% CI 0.78-0.93), where mean diameter was close to chance (0.63, 0.49-0.76, ΔAUROC 0.23, P<0.001). Its value is therefore not confined to examinations a radiologist would likely diagnose anyway. The follow-up configuration shows the same direction of effect (ΔAUROC 0.23, P=0.044), but with only 11 cancers in the ≥1-year stratum its estimates are wide (AUROC 0.78, 95% CI 0.59-0.96).
